# Phenome-wide associations of polygenic scores for schizophrenia and major depression in 100,000 Chinese adults

**DOI:** 10.1101/2025.03.03.25323222

**Authors:** Baihan Wang, Sam Morris, Hannah Fry, Andri Iona, Jonathan Clarke, Kuang Lin, Igor Pupko, Christiana Kartsonaki, Derrick A Bennett, Yiping Chen, Huaidong Du, Ling Yang, Daniel Avery, Dan Schmidt-Valle, Shixian Feng, Dianjianyi Sun, Canqing Yu, Jun Lv, Pei Pei, Junshi Chen, Karoline Kuchenbaecker, Naomi R Wray, Liming Li, Robin G Walters, Zhengming Chen, Iona Y Millwood, China Kadoorie Biobank Collaborative Group

## Abstract

China faces significant mental health challenges, with unique associations between mental disorders and other traits observed in its population. Based on summary statistics in East Asian (EAS) and European (EUR) ancestries, we tested associations of polygenic scores (PGS) for schizophrenia and major depression with 254 phenotypes in 100,640 Chinese adults. PGS predicted schizophrenia (R^2^=1.96%-3.49%) and major depression (R^2^=0.19%-0.77%), and were associated with various socio-demographic, lifestyle, and physical factors. Interestingly, EAS-schizophrenia-PGS was inversely associated with smoking initiation, and EAS-depression-PGS was inversely associated with BMI. Opposing genetic correlations between ancestries were observed for smoking-schizophrenia (inverse in EAS; positive in EUR) and BMI-depression (inverse in EAS; positive in EUR). Mendelian Randomisation supported the causality of these relationships in EUR, but multivariable analyses suggested the influence of pleiotropic effects on other related traits. Our study suggests the context specificity of relationships between mental disorders and other traits, highlighting a potential role of sociocultural factors.

## Introduction

China accounts for 16% of the global burden of mental disorders^1^, creating challenges for China’s national health care system. The number of disability-adjusted life years due to mental disorders in China has been rising steadily, with depressive disorders, anxiety disorders, and schizophrenia (SCZ) contributing to the majority of the burden^2^. However, there is still limited recognition of mental disorders by the general public in China, leading to barriers to diagnosis and treatment.^3^ The resources available for mental health care in China are also considerably lower than those in other high-income countries, with uneven distributions across urban and rural areas^4,5^.

Many mental disorders show significant heritability, with SCZ and major depression (MD) having around 80% and 30% heritability, respectively^6,7^. Breakthroughs in understanding the genetic basis of SCZ and MD have been made by genome-wide association studies (GWAS) in diverse populations^8–12^. The genetic correlations between East Asian ancestry (EAS) and European ancestry (EUR) populations have been estimated to be substantial (> 0.9) for both SCZ and MD^9,10,12^. Nevertheless, important differences still exist between populations. For example, Chen et al. reported larger genetic differences between EAS and EUR for SCZ-associated variants compared to variants randomly selected from the whole genome^13^. Moreover, mental disorders may exhibit distinct characteristics in China. Using Mendelian Randomisation (MR), O’Loughlin et al. reported that lower body mass index (BMI) was causally associated with a higher risk of MD in the Chinese population, which was opposite in direction to the causal association in EUR populations^14^. This suggests that sociocultural factors potentially influence the relationships between mental disorders and other health-related traits^14^.

Polygenic scores (PGS) are the sum of trait-associated variants weighted by their effect sizes and broadly represent an individual’s genetic predisposition for a given trait^15^. They have been found to be predictive of mental disorders in clinical and population-based studies.^16^ PGS can also be employed in phenome-wide association studies (PheWAS), to reveal associations between the genetic risk for mental disorders and a range of other traits across the human phenome.^17^ For example, previous PheWAS conducted in Western and EUR populations reported associations of the PGS for SCZ or MD with other mental disorders, physical illnesses, behavioural traits, and brain structure measures^18–21^, suggesting shared genetic mechanisms and new modifiable risk factors.

Nevertheless, few PheWAS using the PGS for SCZ or MD have been conducted in the Chinese population. Such studies will help discover setting-specific associations, clarify disease aetiology, and inform public health policies. Therefore, the aims of the current study are: 1) to identify associations of the genetic risk for SCZ and MD with 254 phenotypes measured in 100,640 Chinese adults; 2) to examine the consistency of these associations in both EAS and EUR populations with genetic correlation analyses; 3) to assess the causal relevance of these associations using univariable and multivariable MR.

## Results

### PGS associations with schizophrenia and major depression

100,640 genotyped participants in the China Kadoorie Biobank (CKB)^22^ were included in the current study. We computed PGS for SCZ and MD based on two types of GWAS summary statistics (Supplementary Table 1): one from GWAS conducted in EAS (PGS-EAS); the other from both GWAS in EAS and GWAS in EUR (PGS-multi). For SCZ, there were 158 cases recorded during follow-up and 77,148 population-representative controls (Table 1). Both PGS-SCZ-EAS and PGS-SCZ-multi were positively associated with the risk of SCZ. PGS-SCZ-multi explained more variance in SCZ on the liability scale (R^2^ = 3.49%) than PGS-SCZ-EAS (R^2^ = 1.96%). For MD, there were 906 cases (744 recorded at baseline and 169 during follow-up; 7 overlapping) and 77,321 population-representative controls. Similarly, both PGS-MD-EAS and PGS-MD-multi were positively associated with the risk of MD, but PGS-MD-multi explained more variance on the liability scale than PGS-MD-EAS (R^2^ = 0.77% vs 0.19%). The distributions of PGS between cases and controls, as well as OR per PGS quartile are shown in Supplementary Figures 1 and 2. All PGS were associated with their corresponding phenotypes in sex-specific analyses (Supplementary Table 2; Supplementary Figures 3-6.

**Figure 1.**
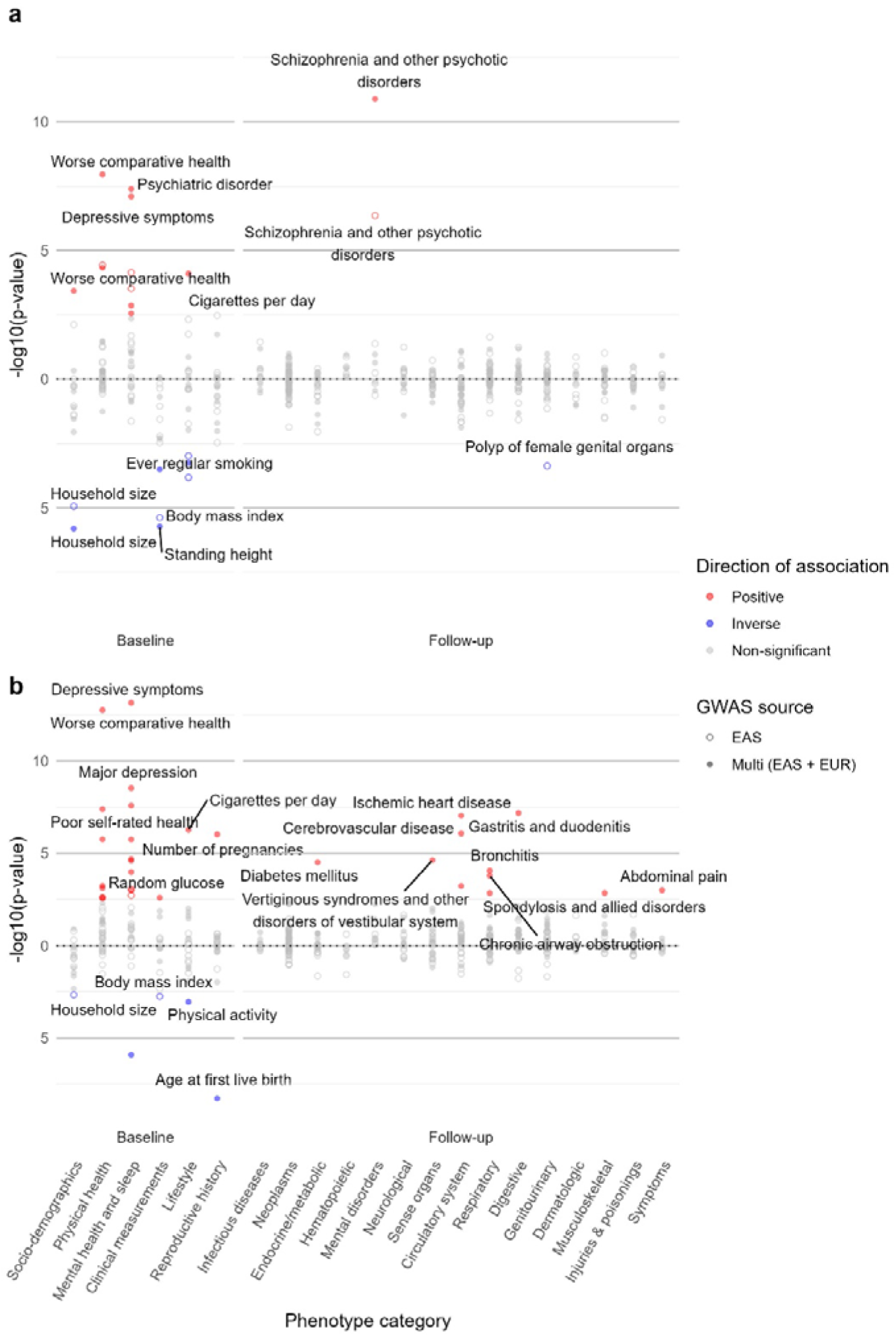
Phenome-wide associations with polygenic scores for schizophrenia and major depression in the China Kadoorie Biobank. Note. a, Results of polygenic scores for schizophrenia. b, Results of polygenic scores for major depression. A total of 254 phenotypes (67 baseline measures and 187 phecodes recorded during follow-up) were tested. Phenotypes only present in one sex are also included and shown here. The shape of dots indicates the GWAS source, while the colour of dots indicates the direction of association. Results were corrected for multiple testing with a false discovery rate = 0.05. The top two most significant associations in each phenotype category are labelled. GWAS: Genome-wide association studies. EAS: East Asian ancestry. EUR: European ancestry.

**Figure 2.**
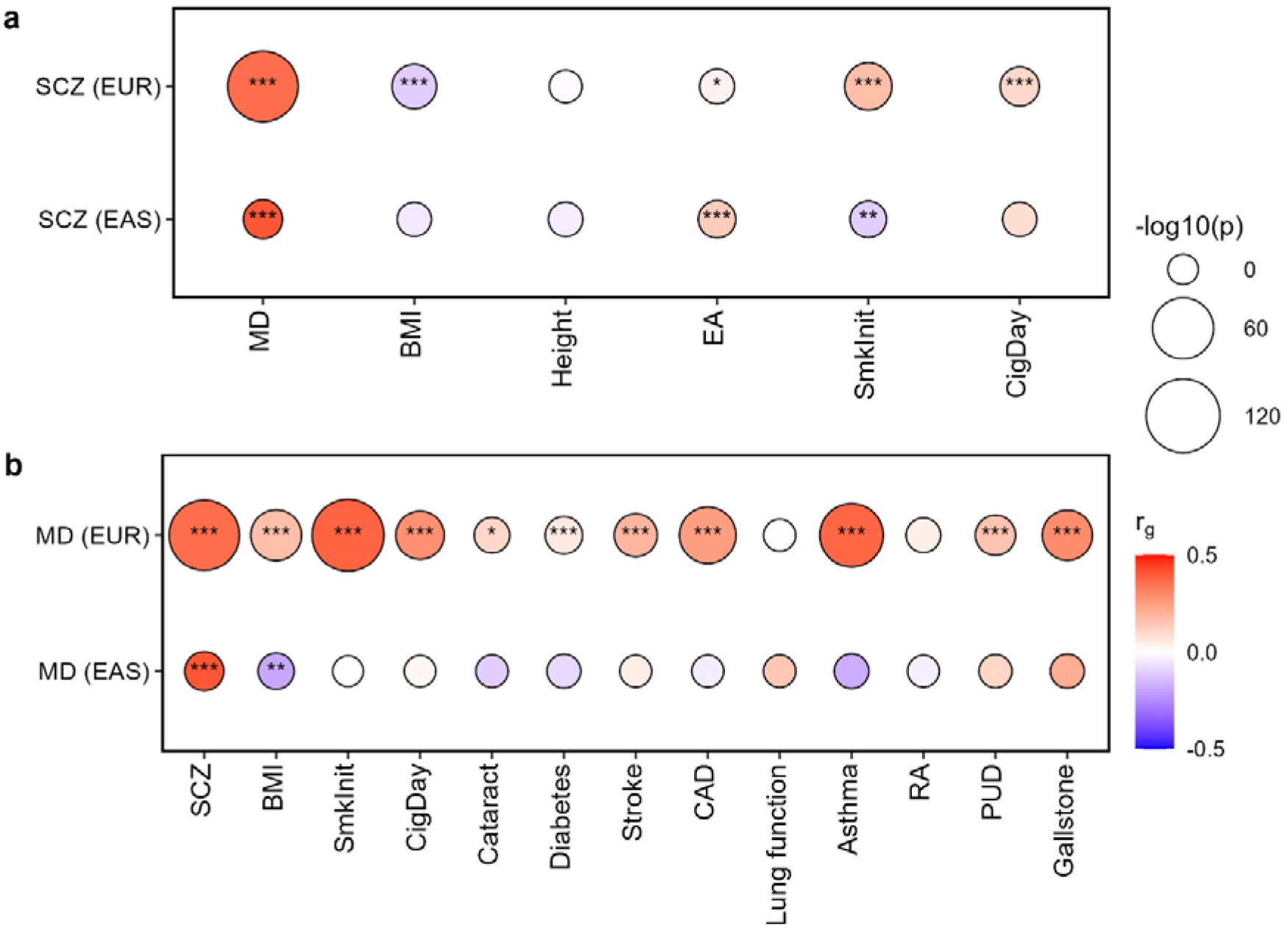
Genetic correlations between schizophrenia/major depression and related phenotypes. Note. a, Genetic correlations with SCZ. b, Genetic correlations with MD. Genetic correlations (r_g_) were tested by LDSC, using GWAS on SCZ/MD and phenotypes within the same ancestry. Mental disorder-phenotype pairs that were significant in the phenome-wide association analysis were tested here. Only phenotypes with publicly available GWAS in both EAS and EUR were included. The colour of the circles indicates the correlation coefficient. The size of the circles is scaled to - log10(*p*-value). *: *p* < 0.05; **: *p* < 0.01; ***: *p* < 0.001. SCZ: Schizophrenia. MD: Major depression. EAS: East Asian ancestry. EUR: European ancestry. BMI: Body mass index. EA: Educational attainment. SmkInit: Smoking initiation. CigDay: Cigarettes per day. CAD: Coronary artery disease. RA: Rheumatoid arthritis. PUD: Peptic ulcer disease.

**Figure 3.**
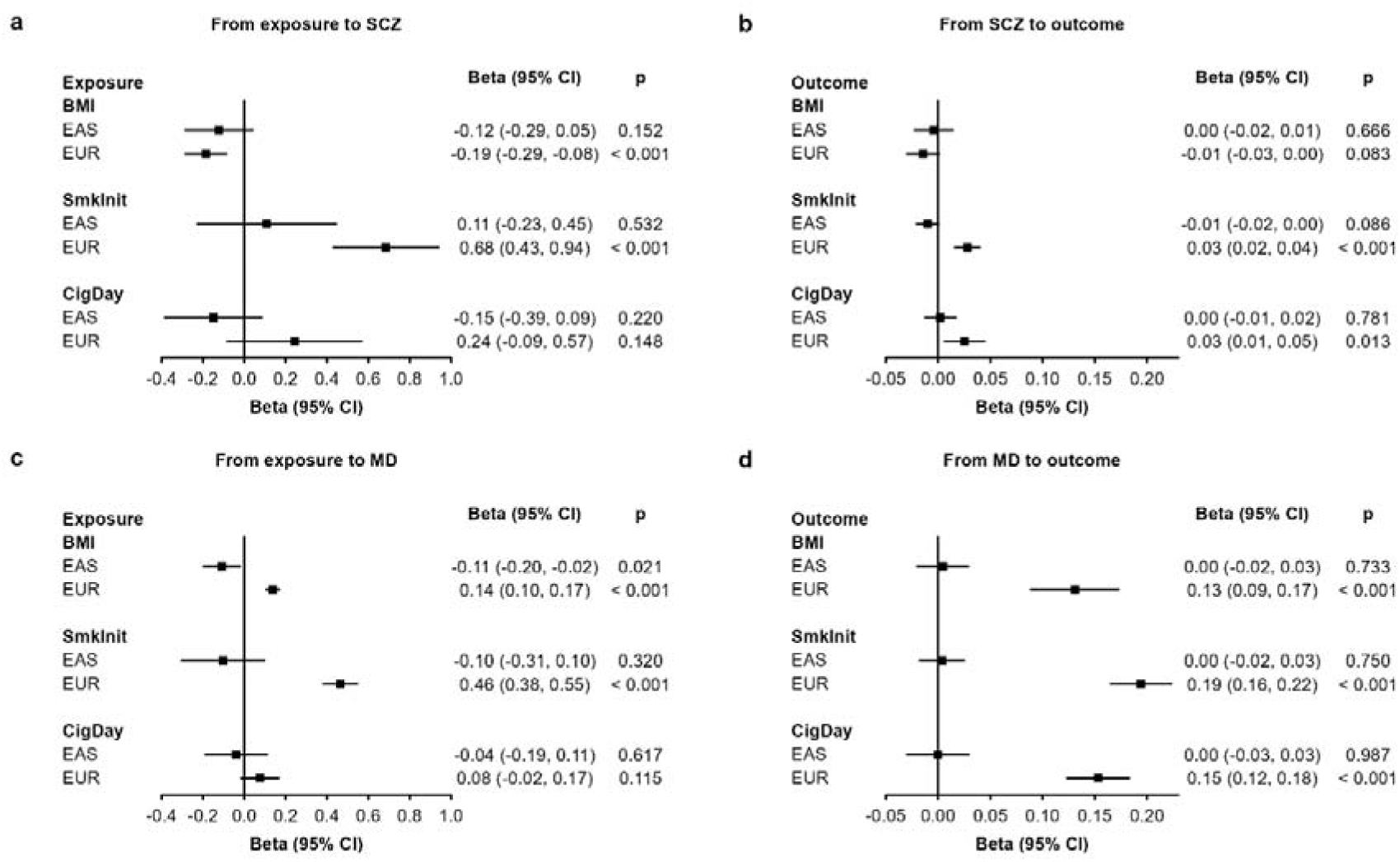
Bi-directional Mendelian Randomisation between schizophrenia/major depression and related phenotypes. Note. a, From exposure to SCZ. b, From SCZ to outcome. c, From exposure to MD. d, From MD to outcome. Results shown here are based on estimates from the inverse variance weighted method. Genetic instruments were selected based on *p* < 1 x 10^−5^ in EAS and *p* < 5 × 10^−8^ in EUR after clumping. SCZ: Schizophrenia. MD: Major depression. EAS: East Asian ancestry. EUR: European ancestry. BMI: Body mass index. SmkInit: Smoking initiation. CigDay: Cigarettes per day. CI: Confidence interval.

**Table 1.** Associations between polygenic scores for schizophrenia/major depression and their corresponding phenotypes in the China Kadoorie Biobank.

| Mental disorder | n (cases) | n (controls) | GWAS source | OR per SD higher PGS (95%CI) | *p* | R<sup>2</sup> on liability scale |
| --- | --- | --- | --- | --- | --- | --- |
| SCZ | 158 | 77,148 | EAS | 1.52 (1.29 - 1.79) | 6.25×10<sup>-7</sup> | 1.96% |
|  |  |  | Multi (EAS + EUR) | 1.73 (1.47 - 2.03) | 3.11×10<sup>-11</sup> | 3.49% |
| MD | 906 | 77,321 | EAS | 1.13 (1.06 - 1.21) | 2.05×10<sup>-4</sup> | 0.19% |
|  |  |  | Multi (EAS + EUR) | 1.27 (1.19 - 1.36) | 8.32×10<sup>-13</sup> | 0.77% |

Note. Cases of mental disorders were identified from the overall dataset, while controls were identified from the population-representative subset. Two types of polygenic scores were assessed for each mental disorder, one based on GWAS in EAS and one based on GWAS in both EAS and EUR. SCZ: Schizophrenia. MD: Major depression. GWAS: Genome-wide association studies. EAS: East Asian ancestry. EUR: European ancestry. OR: Odds ratio per one standard deviation higher polygenic scores. PGS: Polygenic score. CI: Confidence interval.

### Phenome-wide associations

A total of 254 phenotypes (67 at baseline and 197 at follow-up) were included in PheWAS (Figure 1 and Supplementary Data 1). Both PGS-SCZ-EAS and PGS-SCZ-multi were positively associated with a higher risk of psychotic disorders (phecode: 295) during follow-up. At baseline, higher PGS-SCZ-EAS and PGS-SCZ-multi were both associated with self-reported psychiatric disorder and depressive symptoms, as well as worse subjective health and lower household size (*p* < 1 × 10^−4^). A higher PGS-SCZ-EAS was associated with lower BMI (beta per SD higher PGS = −0.06; 95% CI = −0.08 - −0.03; *p* = 4.14 × 10^−6^), while the association between PGS-SCZ-multi and BMI was directionally consistent but did not survive correction for multiple testing (*p* = 0.004). PGS-SCZ-EAS was inversely associated with ever regular smoking (OR = 0.95; 95% CI = 0.93 - 0.98; *p* = 1.53 × 10^−4^) at baseline, while a directionally consistent but non-significant association was found for PGS-SCZ-multi (*p* = 0.069). In contrast, a higher PGS-SCZ-multi was associated with more cigarettes smoked per day, higher levels of education, lower standing/sitting height, and lower physical activity (*p* < 1 × 10^−3^), while directionally consistent associations reaching nominal significance were found for PGS-SCZ-EAS but did not survive for correction for multiple testing (*p* < 0.05). Sex-specific PheWAS showed generally consistent results with the sex-combined analyses (Supplementary Figures 7 and 8; Supplementary Data 1). Adjusting for additional covariates did not substantially change these associations (Supplementary Data 1).

Higher PGS-MD-EAS and PGS-MD-multi were associated with higher risks of self-reported depressive symptoms and MD, as well as worse subjective health at baseline (*p* < 3 × 10^−3^). However, PGS-MD-multi yielded more associations in general than PGS-MD-EAS. At baseline, a higher PGS-MD-multi was associated with more cigarettes smoked per day, shorter and worse sleep, and lower physical activity (*p* < 3 × 10^−3^). A higher PGS-MD-multi was also associated with higher risks of a range of disorders and symptoms, such as anxiety and neurasthenia at baseline, as well as gastritis and duodenitis (phecode: 535), ischemic heart disease (phecode: 411), cerebrovascular disease (phecode: 433), diabetes mellitus (phecode: 250), and chronic airway obstruction (phecode: 496) at follow-up (*p* < 3 × 10^−3^). Conversely, PGS-MD-EAS showed an inverse association with BMI (beta = −0.04, 95% CI = −0.06 - −0.01, *p* = 1.77 × 10^−3^), which was instead positively associated with PGS-MD-multi at nominal significance (beta = 0.03, 95% CI = 0.00 – 0.05, *p* = 0.028). Sex-specific PheWAS yielded generally consistent results with the sex-combined analyses (Supplementary Figures 7 and 8; Supplementary Data 1). For females, we found that higher PGS-MD-multi was associated with a younger age at first birth and a higher number of pregnancies (*p* < 1 × 10^−6^). Adjusting for additional covariates did not substantially change these associations (Supplementary Data 1).

### Genetic correlations

We tested genetic correlations for significant mental disorder-phenotype pairs based on publicly available GWAS summary statistics (Supplementary Table 1). As shown in Figure 2 and Supplementary Table 3, there were positive genetic correlations between SCZ and MD in both EAS and EUR (*p* < 0.001). SCZ had an inverse genetic correlation with smoking initiation in EAS (r_g_ = −0.10; 95% CI = −0.18 - −0.03; *p* = 4.70 × 10^−3^) but a positive genetic correlation with smoking initiation in EUR (r_g_ = 0.17; 95% CI = 0.13 - 0.21; *p* = 6.86 × 10^−19^). We also found positive genetic correlations between SCZ and educational attainment in both EAS and EUR (*p* < 0.05). SCZ had an inverse genetic correlation with BMI and a positive genetic correlation with CigDay in EUR (*p* < 0.001), but neither of these correlations reached statistical significance in EAS. Cross-ancestry analyses showed positive genetic correlations between SCZ and MD, as well as inverse genetic correlations between SCZ and BMI (Supplementary Figure 9 and Supplementary Table 4).

In EAS, there was an inverse genetic correlation between MD and BMI (r_g_ = −0.19; 95% CI = −0.33 - −0.05; *p* = 8.60 × 10^−3^). In contrast, in EUR, MD showed a positive genetic correlation with BMI (r_g_ = 0.17; 95% CI = 0.14 - 0.20; *p* = 1.37 × 10^−26^). We also found positive genetic correlations between MD and smoking initiation, cigarettes per day, and most physical illnesses investigated (*p* < 0.05), but none of these correlations were observed in EAS. Cross-ancestry analyses showed an inverse genetic correlation between MD-EAS and BMI-EUR, as well as positive genetic correlations between MD-EUR and smoking-EAS (Supplementary Figure 9 and Supplementary Table 4).

### Mendelian Randomisation

As BMI, smoking initiation, and cigarettes per day showed associations with SCZ and MD in both PheWAS and genetic correlation analyses, we conducted two-sample bi-directional MR to assess their causal relevance in EAS and EUR. All exposures’ genetic instruments had a mean F-statistic > 20, indicating low weak instrument bias (Supplementary Data 2). In EAS, there was an inverse association between genetically predicted BMI and MD (beta = −0.11; 95% CI = −0.20 - −0.02; *p* = 0.021; Figure 3), but this association was no longer significant in our sensitivity analyses with alternative MR methods or with a more stringent *p*-value threshold (Supplementary Data 2 and Supplementary Figure 10). In EUR, genetically predicted BMI was inversely associated with SCZ (beta = −0.19; 95% CI = −0.29 - −0.08; *p* < 0.001) but positively associated with MD (beta = 0.14; 95% CI = 0.10 - 0.17; *p* < 0.001). There were also strong positive bi-directional causal associations between smoking initiation and both SCZ and MD in EUR (*p* < 0.001). Additionally, genetically predicted SCZ and MD were both positively associated with cigarettes per day in EUR (*p* < 0.05). Sensitivity analyses using other MR methods showed consistent results in EUR (Supplementary Data 2).

We performed multivariable MR to assess if the causal associations observed in EUR could be influenced by pleiotropic effects on other related traits. Compared to univariable MR results, adding educational attainment as an additional exposure attenuated the causal relationship of BMI on SCZ (Figure 4, Supplementary Data 3). The causal relevance of MD to BMI was also attenuated when including smoking initiation as an additional exposure. Furthermore, we found that the bi-directional association between smoking initiation and SCZ was attenuated by adding cannabis use disorder as an additional exposure. The conditional F-statistics were > 10 for most exposures, but all models had a *p* < 0.05 for their Q-statistics, indicating potential pleiotropy. We then used Q-statistic minimisation to re-estimate betas to account for weak instruments and pleiotropy, which shifted most of the estimates closer to 0 (Supplementary Data 3).

**Figure 4.**
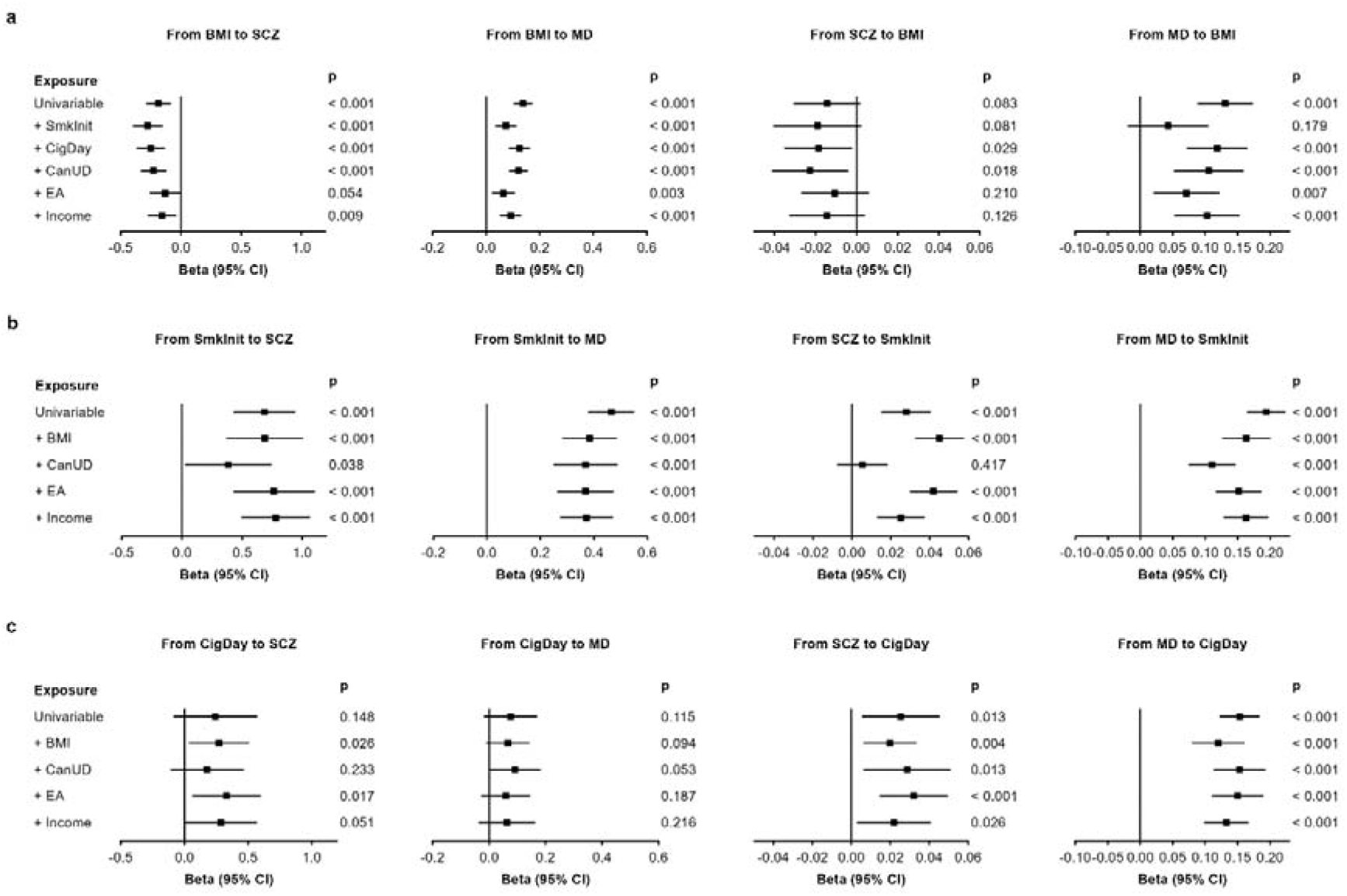
Multivariable Mendelian Randomisation between schizophrenia/major depression and other phenotypes in the European ancestry population. Note. a, Results for BMI. b, Results for SmkInit. c, Results for CigDay. All multivariable Mendelian Randomisation analyses were based on GWAS summary statistics in the European ancestry population with the inverse variance weighted method. Genetic instruments were selected based on *p* < 5 × 10^−8^ after clumping. An additional phenotype was added to the univariable model to assess its influence on the association each time. SCZ: Schizophrenia. MD: Major depression. BMI: Body mass index. SmkInit: Smoking initiation. CigDay: Cigarettes per day. CanUD: Cannabis use disorder. EA: Educational attainment. CI: Confidence interval.

## Discussion

This is the first study to systematically investigate the phenome-wide associations of PGS for SCZ and MD in the Chinese population. Both PGS were associated with their corresponding mental disorders, as well as with a range of socio-demographic, lifestyle, and physical measures. In particular, we identified population-specific inverse associations between genetic susceptibility for SCZ and smoking initiation, and between genetic susceptibility for MD and BMI. Genetic correlation analyses revealed opposing findings for these associations in EAS and EUR, although MR only supported causal relationships in EUR. Further multivariable MR analyses suggested that such causality in EUR might be accounted for by pleiotropic effects on other related traits.

Many of our PheWAS results in this Chinese population replicated previous findings in Western populations. For example, the PGS for SCZ and MD were associated with various mental-health-related phenotypes in the current study, agreeing with the considerable genetic correlations between different mental disorders in EUR^23^. Notably, both PGS-SCZ and PGS-MD were strongly associated with self-reported subjective health measures, which might reflect the high prevalence of somatic symptoms in patients with mental disorders in China^24^. Moreover, we found that PGS-SCZ was inversely associated with BMI and positively associated with educational level, replicating previous findings of genetic analyses in EUR^25,26^. Additionally, we found that a higher PGS-MD-multi was associated with a younger age at first birth and a higher number of pregnancies in Chinese women, consistent with previous observational findings in the US population^27,28^. PGS-MD-multi was also associated with the risks of several physical illnesses at follow-up in CKB, supporting previous observational associations of MD with ischaemic heart disease^29^, stroke^30^, COPD^31^, and diabetes^32,33^ in diverse populations. Nevertheless, the associations of PGS-MD-multi with reproductive factors and physical illnesses were not found for PGS-MD-EAS, nor in the following genetic correlation analyses in EAS. This might be due to the limited power of GWAS on MD in EAS, or could suggest potentially different genetic architecture of MD between EAS and EUR.

In PheWAS, we observed a unique inverse association between PGS-MD-EAS and BMI, as well as an inverse genetic correlation between MD and BMI in EAS. Conversely, a positive genetic correlation between BMI and MD in EUR was observed in the current and previous studies^34^. Further MR analyses revealed a positive association between genetically predicted BMI and MD in EUR but an inverse association in EAS, although the latter was not significant in our sensitivity analyses. The results generally agree with the inverse association between genetically predicted BMI and MD in EAS reported by O’Loughlin et al^14^. The weak association between genetically predicted BMI and MD in EAS in our study might be explained by the limited power of GWAS on MD and BMI in EAS. However, considering their inverse association in PheWAS and inverse genetic correlation in EAS, it is unlikely that an increase in GWAS sample size would change the direction of this association in MR. Differences in sociocultural factors may offer an alternative explanation: It has been hypothesised that higher BMI is viewed as a sign of wealth and health among middle-aged and old adults in China, but people with higher BMI face more weight-based discrimination in Western societies^35^. Additionally, the maladaptive coping explanation postulates that the positive association between MD and BMI might be due to the engagement in unhealthy behaviours among people with MD^36^, and such coping strategies might be more commonly adopted in Western societies^37^. Consistent with this explanation, our MVMR analyses revealed that the causal relationship from MD to BMI in EUR was attenuated after adding smoking initiation as an additional exposure. The result also agrees with a previous observational study in the Belgian population, where lifestyle factors (e.g. smoking, physical activity, and disordered eating) mediated the association between poor mental health and excess weight^38^.

In PheWAS, PGS-SCZ-EAS was inversely associated with smoking initiation, which was supported by the inverse genetic correlation between SCZ and smoking initiation in EAS. Conversely, both smoking initiation and cigarettes per day showed positive genetic correlations with SCZ in EUR. Previous research in Western populations reported a positive association between smoking and SCZ^39^, with MR studies suggesting a bi-directional causal relationship in EUR^40^. Hence, the self-medication hypothesis postulates that smoking may act as a way to relieve symptoms among people with schizophrenia^41^. However, studies in China reported a similar prevalence of smoking in people with schizophrenia to the prevalence in the general population^42,43^. Our MR analysis in EAS also revealed a lack of causal associations between SCZ and smoking initiation. Such differences between populations may be explained by differences in smoking behaviour: while there were fewer current smokers (11.9%) than ex-smokers (24.9%) in the UK^44^, the proportion of current smokers (24.3%) was much higher than that of ex-smokers (3.1%) in China^45^. This is also related to the fact that smoking in China is often accepted as a social activity, especially for men^46^. Therefore, it is possible that people with higher PGS-SCZ engage less in social activities, and thus are less likely to smoke.

Cannabis use is another possible explanation for this difference, as it is a strong risk factor for SCZ^47^ and often co-occurs with cigarette smoking in Western populations^48^. In contrast, very few people use cannabis in China due to strict regulations, so the relationship between smoking and SCZ would not be influenced by cannabis use. Indeed, in our multivariable MR analysis, adding cannabis use disorder as an additional exposure attenuated the causal associations between smoking initiation and SCZ in EUR. This suggests that the causality of smoking initiation and SCZ might be, at least partially, explained by pleiotropic effects on cannabis use.

The current study has limitations. First, since the CKB follow-up data came from hospitalisation and death records, certain medical conditions might have been under-represented, such as episodes of mental disorders that did not require inpatient treatment. Moreover, although we computed PGS based on the largest GWAS available at the time of the study, GWAS in EAS are still less powered than GWAS in EUR, and there was currently no well-powered GWAS on bipolar disorder in EAS. Nevertheless, all PGS in the current study were still strongly associated with SCZ and MD, supporting their validity. Additionally, although we employed MR for causal inference, we were unable to do this for certain phenotypes (e.g. sleep duration) due to the lack of publicly available GWAS in EAS. Furthermore, although all exposures’ genetic instruments had a mean F-statistic > 20 in the univariable MR analyses, some of them (e.g. cannabis use disorder) had a conditional F-statistic < 10 in the multivariable MR analyses. However, since Q-statistic minimisation in the sensitivity analyses generally shifted the betas towards the null, we believe that the conclusions drawn from the multivariable MR are still valid. Finally, some of our PheWAS findings still require further explanation, as it is unclear why PGS-SCZ-EAS was inversely associated with female genital polyps. Although oestrogen deficiency has been associated with SCZ^49^, more research is needed to replicate this association in other cohorts.

To conclude, the current study found that the PGS for SCZ and MD were associated with a range of phenotypes among ∼100,000 Chinese adults, suggesting their shared genetic architectures. The distinct associations of smoking-SCZ and BMI-MD across different populations highlight the important role sociocultural factors may play in those relationships. More research in diverse populations is still needed to clarify the context specificity of mental disorders and their links with other health-related traits. Policymakers should also be aware of such differences to develop better mental health strategies tailored for specific populations.

## Methods

### Study population

The China Kadoorie Biobank (CKB) is a prospective cohort study with > 512,000 adults aged 30–79 years, recruited during 2004-08 from 10 regions across urban and rural areas in China^22^. At baseline, participants were assessed by laptop-based questionnaires on socio-demographic characteristics, medical history, and lifestyle habits. Physical measurements (e.g. anthropometry, blood pressure, and heart rate) were also collected at baseline. After the baseline survey, the long-term health of the participants was monitored via linkage with local death or disease registries, as well as the national health insurance systems recording all hospitalisation episodes. The present study is based on follow-up to 1^st^ January 2019 (median = 12.36 [Q1 – Q3: 11.20 – 13.27] years follow-up).

CKB complies with all required ethical standards for medical research on human subjects. Ethical approval was obtained from the Ethical Review Committee of the Chinese Centre for Disease Control and Prevention (Beijing, China, 005/2004) and the Oxford Tropical Research Ethics Committee, University of Oxford (UK, 025-04). All participants provided written informed consent.

### Genotyping

A total of 100,640 CKB participants were genotyped using a custom Affymetrix array and included in the current study^50^. Of these participants, 23,518 were cases of cardiovascular disease (CVD) or chronic obstructive pulmonary disease (COPD) selected for a nested case-control study, while the remaining 77,122 were randomly selected and population-representative. Details of genotyping and quality control procedures in CKB have been previously described^50^. Genotyped variants were pre-phased using SHAPEIT v4.2 (SHAPEIT v2.904 for chromosome X)^51^ and uploaded to the TOPMed^52^ or Westlake Biobank for Chinese^53^ server for imputation. Two sets of the imputed data were merged by selecting the imputed genotype with a higher imputation INFO score for each variant.

### Polygenic score computation

We computed PGS for SCZ and MD based on two types of GWAS summary statistics: one from GWAS conducted in EAS; the other from both GWAS in EAS and GWAS in EUR. This is because variants discovered by GWAS in EAS may include population- and setting-specific genetic associations, while incorporating GWAS in multi-ancestries has been found to improve the overall predictive power of PGS^54^. As a result, four different PGS were computed for each participant in CKB: PGS-SCZ-EAS, PGS-SCZ-multi (EAS+EUR), PGS-MD-EAS, and PGS-MD-multi (EAS+EUR).

GWAS summary statistics for SCZ^8,10^ and MD^9,11^ in EAS and EUR were downloaded from the Psychiatric Genomics Consortium (PGC) website (https://pgc.unc.edu/). Since CKB contributed to the GWAS on MD in EAS, the GWAS summary statistics excluding CKB data were used here to ensure no sample overlap. We used PRS-CSx^54,55^ to infer the posterior SNP effect sizes of variants via a continuous shrinkage prior within EAS, or via a shared prior across EAS and EUR. We used samples from the EAS and EUR superpopulations in the 1000 Genomes Project^56^ as linkage disequilibrium (LD) references. Only HapMap3 variants with a minor allele frequency (MAF) > 0.01 in the relevant ancestry were included in the analysis, as recommended by PRS-CSx^54,55,57^.

Based on the SNP effect sizes generated by PRS-CSx, we then used PLINK^58^ to compute PGS for all 100,640 genotyped participants in CKB. The PGS were then standardised based on the means and SDs among the 77,122 population-representative CKB participants.

### Associations with schizophrenia and major depression

We first tested the associations between the newly computed PGS and their corresponding phenotypes (SCZ and MD) in CKB. Cases of SCZ were identified based on hospitalisation records at follow-up (ICD-10 code: F25). Cases of MD were identified based on both the Composite International Diagnostic Inventory-short form (CIDI-SF) completed at baseline, and hospitalisation records at follow-up (ICD-10 codes: F32, F33, F34.1, and F38.1). To maximise power and avoid potential biases, we identified cases from the overall dataset (including both the selected CVD/COPD cases and the population-representative participants), while controls were only identified from the population-representative subset (Supplementary Figure 11). With SCZ/MD as the outcome variable, we ran logistic regression analyses with the standardised PGS for SCZ/MD as the main explanatory variable, including age, age^2^, sex, study region, and the first 11 genomic principal components as covariates. To assess associations in different risk groups, the PGS were further split into quartiles to compute odds ratios with the first quartile as the reference.

### Phenome-wide association studies

We included 254 phenotypes measured at baseline or during follow-up in PheWAS. These included 67 baseline characteristics capturing socio-demographics, physical health, mental health and sleep, clinical measurements, lifestyle, and female reproductive history. Categorical variables with more than two levels were converted to binary variables, as indicated by their names in Supplementary Data 1 and 2. Hospitalisation episodes for diseases and conditions at follow-up were originally recorded as ICD-10 codes. We then used phecodes (version 1.2)^59^ to curate the ICD-10 codes and generate 187 clinically defined phecodes in CKB. To maximise power, we analysed the three-digit parent codes in PheWAS and only kept phecodes with > 100 cases. This resulted in 80% power to detect an OR of 1.5 per SD higher PGS at *p* < 0.001.

We applied logistic and linear regression models to assess the associations between PGS and categorical and continuous phenotypes, respectively, in CKB. Age, age^2^, sex, study region, and the first 11 genomic principal components were included as covariates. To maximise power and avoid potential biases, we identified cases of all disease-related phenotypes from the overall dataset (including both the selected CVD/COPD cases and the population-representative participants), while controls were only identified from the population-representative subset (Supplementary Figure 11). For each phecode, we excluded controls with related conditions, as defined by its phecode exclude range^59^. Analyses of all other phenotypes were restricted to the population-representative participants. Multiple testing was corrected with a false discovery rate = 0.05. The same procedure was repeated in sex-specific analyses, for all phenotypes were included in the overall analysis. For associations identified in PheWAS, we performed sensitivity analyses by including household size, household income, ownership index, education level, BMI, smoking, and alcohol drinking as additional covariates.

### Genetic correlations

We estimated the genetic correlations, based on GWAS summary statistics, for mental disorder PGS-phenotype pairs that showed associations in PheWAS. This was to examine if the findings in CKB were generalizable to the larger EAS population, and if the associations were consistent between EAS and EUR. We downloaded the GWAS summary statistics for those phenotypes from published meta-analyses, genomics consortia, or other cohorts/biobanks, if they were publicly available in both EAS and EUR. For GWAS in EAS, we selected studies that were not conducted in CKB, or meta-analyses that included CKB as only one of the many contributing cohorts. These GWAS summary statistics are summarised in Supplementary Table 1. All GWAS summary statistics were processed and harmonised with the GWASLab package^60^.

We used LDSC^61^ to compute the genetic correlations (r_g_) for all mental disorder-phenotype pairs within each ancestry. We also performed additional analyses using Popcorn^62^ to compute the genetic-effect correlations (ρ_ge_) across EAS and EUR. All genetic correlation analyses were conducted with variants with MAF > 0.01 and present in HapMap3^57^, based on EAS and EUR LD references from the 1000 Genomes Project^56^.

### Mendelian Randomisation

For mental disorder-phenotype pairs that showed associations in both PheWAS and genetic correlation analyses, we used MR to assess their causal relevance. There are three core assumptions of MR: 1) the genetic variants are associated with the exposure (relevance assumption); 2) there are no unmeasured confounders between the genetic variants and the outcome (independence assumption); 3) the genetic variants do not affect the outcome expect via the exposure (exclusion restriction assumption).

We conducted bi-directional two-sample MR using the TwoSampleMR^63^ package. To maximise power and distinguish MR from PheWAS, we performed summary-level MR based on publicly available GWAS summary statistics instead of individual-level MR. To assess MR assumption 1), we calculated the mean F-statistics for each exposure to assess instrument strength. To meet MR assumption 2), we performed MR with exposure and outcome GWAS from the same ancestry, separately for EAS and EUR, to avoid confounding due to ancestry. To remove correlated variants, we performed LD clumping with window = 10000 kb and r^2^ = 0.001 based on EAS/EUR LD references from the 1000 Genomes Project^56^. Due to the relatively small sample sizes of GWAS in EAS, we selected variants that reached suggestive genome-wide significance (*p* = 1 x 10^−5^) as genetic instruments in the main analyses^64^. We also performed sensitivity analyses with variants that reached genome-wide significance (*p* = 5 × 10^−8^) as genetic instruments in EAS. Genetic instruments in EUR were selected only if they reached genome-wide significance. We used four different methods to perform the two-sample MR: inverse-variance weighting (IVW; main method)^64^; MR-Egger^65^; weighted median^66^; weighted mode^67^.

To assess MR assumption 3), we further performed summary-level multivariable MR^68^ using the TwoSampleMR^63^ and MVMR^69^ packages to account for potential pleiotropic effects on other phenotypes (i.e. BMI, smoking, educational attainment, and income) in EUR. This was done by adding each of the extra phenotypes as an additional exposure, and then comparing the multivariable MR results with the univariable MR results. To conduct multivariable MR, we first performed LD clumping for all pairs of exposures with window = 10000 kb, r^2^ = 0.001, and *p* = 5 × 10^−8^, based on the EUR LD reference from the 1000 Genomes Project^56^. We then estimated the pairwise covariances between each instrument and its corresponding pairs of exposures, based on the phenotypic correlations between exposures in the UK Biobank (Supplementary Note 2; Supplementary Table 5). We also estimated conditional F-statistics to test instrument strength and Q-statistics to test horizontal pleiotropy^68^. The main multivariable MR analysis was conducted using an IVW model, taking into account the covariance matrix between exposures. Additionally, we used Q-statistic minimisation to re-estimate beta, which is robust to weak instruments and pleiotropy^68^. All univariable and multivariable MR analyses were conducted in R-4.4.1^70^.

## Data Availability

In CKB, non-genetic data (e.g. baseline, resurveys, biomarkers, and disease endpoints) are available and updated periodically for access by bona fide researchers. Details of the CKB Data Sharing Policy, data release schedules and data request application procedures are available at www.ckbiobank.org. All queries about data access can be made to. Access to individual participant genetic data (e.g. genotyping, whole genome sequence) is currently constrained by China's Administrative Regulations on Human Genetic Resources, for which collaboration with CKB researchers is generally required, which may be subject to separate regulatory approvals in China if it involves substantial sharing of unpublished data. Further information is available from the corresponding authors upon request. Summary statistics of genome-wide association studies used in this study are publicly available, as summarised in Supplementary Table 1.

## Acknowledgement

The chief acknowledgement is to the participants, the project staff, and the China National Centre for Disease Control and Prevention (CDC) and its regional offices for assisting with the fieldwork. We thank Judith Mackay in Hong Kong; Yu Wang, Gonghuan Yang, Zhengfu Qiang, Lin Feng, Maigeng Zhou, Wenhua Zhao, Yan Zhang in China CDC; Lingzhi Kong, Xiucheng Yu, and Kun Li in the Chinese Ministry of Health; and Garry Lancaster, Sarah Clark, Martin Radley, Mike Hill, Hongchao Pan, and Jill Boreham in the CTSU, Oxford, for assisting with the design, planning, organization, and conduct of the study.

The CKB baseline survey and the first re-survey were supported by the Kadoorie Charitable Foundation in Hong Kong. The long-term follow-up has been supported by Wellcome grants to Oxford University (212946/Z/18/Z, 202922/Z/16/Z, 104085/Z/14/Z, 088158/Z/09/Z) and grants from the Noncommunicable Chronic Diseases-National Science and Technology Major Project (2023ZD0510100), and the National Natural Science Foundation of China (82192900, 82192901, 82192904, 82388102). The UK Medical Research Council (MC_UU_00017/1, MC_UU_12026/2, MC_U137686851), Cancer Research UK (C16077/A29186; C500/A16896) and the British Heart Foundation (CH/1996001/9454), provide core funding to the Clinical Trial Service Unit and Epidemiological Studies Unit at Oxford University for the project. DNA extraction and genotyping were supported by GlaxoSmithKline and the UK Medical Research Council (MC-PC-13049, MC-PC-14135). Computation used the Oxford Biomedical Research Computing (BMRC) facility, a joint development between the Wellcome Centre for Human Genetics and the Big Data Institute supported by Health Data Research UK and the NIHR Oxford Biomedical Research Centre. The views expressed are those of the author(s) and not necessarily those of the NHS, the NIHR or the Department of Health. Analyses in the UK Biobank have been conducted using the UK Biobank Resource under Application Number 50474. Baihan Wang is funded by the Nuffield Department of Population Health Early Career Research Fellowship.

## Data availability

In CKB, non-genetic data (e.g. baseline, resurveys, biomarkers, and disease endpoints) are available and updated periodically for access by bona fide researchers. Details of the CKB Data Sharing Policy, data release schedules and data request application procedures are available at www.ckbbiobank.org. Accessing individual participant genetic data (e.g. genotyping, whole genome sequence) is currently constrained by China’s Administrative Regulations on Human Genetic Resources, for which collaboration with CKB researchers is generally required, subject to separate approvals. Further information is available from the corresponding authors upon request. Summary statistics of genome-wide association studies used in this study are publicly available, as summarised in Supplementary Table 1.

