## Supplementary for "Phenome-wide associations of polygenic scores for schizophrenia and major depression in 100,000 Chinese adults"

### Supplementary Note 1. Members of CKB Collaborative Group

**International Steering Committee:** Junshi Chen, Zhengming Chen (PI), Robert Clarke, Rory Collins, Liming Li (PI), Jun Lv, Richard Peto, Robin Walters.

**International Co-ordinating Centre, Oxford:** Daniel Avery, Maxim Barnard, Derrick Bennett, Ruth Boxall, Ka Hung Chan, Yiping Chen, Zhengming Chen, Charlotte Clarke, Jonathan Clarke; Robert Clarke, Huaidong Du, Ahmed Edris Mohamed, Hannah Fry, Simon Gilbert, Prapthi Harish, Pek Kei Im, Andri Iona, Maria Kakkoura, Christiana Kartsonaki, Kshitij Kolhe, Hubert Lam, Kuang Lin, James Liu, Mohsen Mazidi, Iona Millwood, Sam Morris, Qunhua Nie, Alfred Pozarickij, Maryam Rahmati, Paul Ryder, Dan Schmidt, Becky Stevens, Iain Turnbull, Robin Walters, Baihan Wang, Lin Wang, Neil Wright, Ling Yang, Xiaoming Yang, Pang Yao.

**National Co-ordinating Centre, Beijing:** Xiao Han, Can Hou, Qingmei Xia, Chao Liu, Jun Lv, Pei Pei, Dianjianyi Sun, Canqing Yu, Lang Pan.

**10 Regional Co-ordinating Centres:**

**Qingdao CDC:** Zengchang Pang, Ruqin Gao, Shanpeng Li, Haiping Duan, Shaojie Wang, Yongmei Liu, Ranran Du, Yajing Zang, Liang Cheng, Xiaocao Tian, Hua Zhang, Yaoming Zhai, Feng Ning, Xiaohui Sun, Feifei Li. **Licang CDC:** Silu Lv, Junzheng Wang, Wei Hou. **Heilongjiang Provincial CDC:** Wei Sun, Shichun Yan, Xiaoming Cui. **Nangang CDC:** Chi Wang, Zhenyuan Wu, Yanjie Li, Quan Kang. **Hainan Provincial CDC:** Huiming Luo, Tingting Ou. **Meilan CDC:** Xiangyang Zheng, Zhendong Guo, Shukuan Wu, Yilei Li, Huimei Li. **Jiangsu Provincial CDC:** Ming Wu, Yonglin Zhou, Jinyi Zhou, Ran Tao, Jie Yang, Jian Su. **Suzhou CDC:** Fang Liu, Jun Zhang, Yihe Hu, Yan Lu, Liangcai Ma, Aiyu Tang, Shuo Zhang, Jianrong Jin, Jingchao Liu. **Guangxi Provincial CDC:** Mei Lin, Zhenzhen Lu. **Liuzhou CDC:** Lifang Zhou, Changping Xie, Jian Lan, Tingping Zhu, Yun Liu, Liuping Wei, Liyuan Zhou, Ningyu Chen, Yulu Qin, Sisi Wang. **Sichuan Provincial CDC:** Xianping Wu, Ningmei Zhang, Xiaofang Chen, Xiaoyu Chang. **Pengzhou CDC:** Mingqiang Yuan, Xia Wu, Xiaofang Chen, Wei Jiang, Jiaqiu Liu, Qiang Sun. **Gansu Provincial CDC:** Faqing Chen, Xiaolan Ren, Caixia Dong. **Maiji CDC:** Hui Zhang, Enke Mao, Xiaoping Wang, Tao Wang, Xi zhang. **Henan Provincial CDC:** Kai Kang, Shixian Feng, Huizi Tian, Lei Fan. **Huixian CDC:** XiaoLin Li, Huarong Sun, Pan He, Xukui Zhang. **Zhejiang Provincial CDC:** Min Yu, Ruying Hu, Hao Wang. **Tongxiang CDC**: Xiaoyi Zhang, Yuan Cao, Kaixu Xie, Lingli Chen, Dun Shen. **Hunan Provincial CDC:** Xiaojun Li, Donghui Jin, Li Yin, Huilin Liu, Zhongxi Fu. **Liuyang CDC:** Xin Xu, Hao Zhang, Jianwei Chen, Yuan Peng, Libo Zhang, Chan Qu.

### Supplementary Note 2. Estimation of covariance matrix in multivariable Mendelian Randomisation.

A covariance matrix containing pairwise covariances between an instrument and pairs of exposures is required for causal effect estimation and sensitivity analyses in multivariable MR. This matrix can be estimated based on the phenotypic correlations between exposures using the MVMR package in R.

We used the UK Biobank data to calculate the pairwise phenotypic correlations between BMI, smoking initiation (ex- + current smokers), cigarettes per day (among current smokers), heavy cannabis use (lifetime cannabis use > 100 times), age at completion of full time education, average annual household income, schizophrenia, and major depression. As many exposures were not continuous, we used Spearman’s rank-based correlation to compute the correlation coefficient rho. Since very few people in UKB had cannabis use disorder, we used heavy cannabis use as a proxy. Schizophrenia and major depression were defined based on self-reported diagnosis at baseline, as well as hospitalisation and death records at follow-up. All analyses were restricted to participants with self-reported White ethnicity. The correlation coefficients are shown in Supplementary Table 5.

### Supplementary Table 1. GWAS summary statistics used in analyses.

| **Phenotype** | **Ancestry** | **Source** | **First author (year)** | **PMID/URL** |  |
| --- | --- | --- | --- | --- | --- |
| Schizophrenia | EAS | PGC | Trubetskoy (2022) | 35396580 |  |
| Schizophrenia | EUR | PGC | Trubetskoy (2022) | 35396580 |  |
| Major depression | EAS | PGC | Meng (2024) | 38177345 |  |
| Major depression | EUR | PGC | Adams (2025) | 39814019 |  |
| Body mass index | EAS | BBJ | Akiyama (2017) | 28892062 |  |
| Body mass index | EUR | GIANT | Yengo (2018) | 30124842 |  |
| Height | EAS | GIANT | Yengo (2022) | 36224396 |  |
| Height | EUR | GIANT | Yengo (2022) | 36224396 |  |
| Smoking initiation | EAS | GSCAN | Saunders (2022) | 36477530 |  |
| Smoking initiation | EUR | GSCAN | Saunders (2022) | 36477530 |  |
| Cigarettes per day | EAS | GSCAN | Saunders (2022) | 36477530 |  |
| Cigarettes per day | EUR | GSCAN | Saunders (2022) | 36477530 |  |
| Cataract | EAS | BBJ | Sakaue (2021) | 34594039 |  |
| Cataract | EUR | UKB | The Neale Lab (2018) | <http://www.nealelab.is/uk-biobank/> |  |
| Diabetes | EAS | Meta-analysis | Suzuki (2024) | 38374256 |  |
| Diabetes | EUR | Meta-analysis | Suzuki (2024) | 38374256 |  |
| Stroke | EAS | GIGASTROKE | Mishra (2022) | 36180795 |  |
| Stroke | EUR | GIGASTROKE | Mishra (2022) | 36180795 |  |
| Coronary artery disease | EAS | BBJ | Koyama (2020) | 33020668 |  |
| Coronary artery disease | EUR | CARDIoGRAMplusC4D | Aragam (2022) | 36474045 |  |
| Lung function (FEV1/FVC) | EAS | TWB | Chen (2023) | 38116116 |  |
| Lung function (FEV1/FVC) | EUR | Meta-analysis | Shrine (2023) | 36914875 | 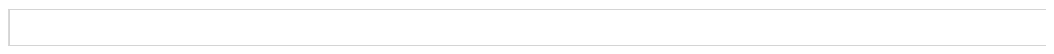 |
| Asthma | EAS | GBMI | Tsuo (2022) | 36778051 |  |
| Asthma | EUR | GBMI | Tsuo (2022) | 36778051 |  |
| Rheumatoid arthritis | EAS | Meta-analysis | Ishigaki (2022) | 36333501 |  |
| Rheumatoid arthritis | EUR | Meta-analysis | Ishigaki (2022) | 36333501 |  |
| Pectic ulcer disease | EAS | Meta-analysis | He (2023) | 38036781 |  |
| Pectic ulcer disease | EUR | UKB | Wu (2021) | 33608531 |  |
| Gallstone | EAS | BBJ | Sakaue (2021) | 34594039 |  |
| Gallstone | EUR | Meta-analysis | Fairfield (2022) | 34651315 |  |
| Educational attainment | EUR | SSGAC | Okbay (2022) | 35361970 |  |
| Income | EUR | Meta-analysis | Kweon (2025) | 39875632 |  |

Note. EAS: East Asian ancestry. EUR: European ancestry. BBJ: Biobank Japan. UKB: UK Biobank. TWB: Taiwan Biobank.

### Supplementary Table 2. Associations between polygenic scores for schizophrenia/major depression and their corresponding phenotypes in CKB by sex.

| **Sex** | **Mental disorder** | **n (cases)** | **n (controls)** | **GWAS source** | **OR per SD higher PGS (95% CI)** | ***p*** | **R2 on liability scale** |
| --- | --- | --- | --- | --- | --- | --- | --- |
| Female | SCZ | 95 | 45,717 | EAS | 1.55 (1.26 - 1.92) | 4.70×10^-5^ | 2.29% |
|  |  |  |  | Multi  (EAS + EUR) | 1.87 (1.52 - 2.31) | 4.05×10^-9^ | 4.83% |
|  | MD | 621 | 45,296 | EAS | 1.11 (1.02 - 1.20) | 0.014 | 0.11% |
|  |  |  |  | Multi  (EAS + EUR) | 1.28 (1.19 - 1.39) | 9.40×10^-10^ | 0.80% |
| Male | SCZ | 63 | 31,273 | EAS | 1.46 (1.13 - 1.89) | 0.004 | 1.51% |
|  |  |  |  | Multi  (EAS + EUR) | 1.52 (1.18 - 1.96) | 0.001 | 1.89% |
|  | MD | 285 | 31,119 | EAS | 1.19 (1.06 - 1.34) | 0.003 | 0.44% |
|  |  |  |  | Multi  (EAS + EUR) | 1.25 (1.11 - 1.41) | 1.79×10^-4^ | 0.65% |

Note. Cases of mental disorders were identified from the overall dataset, while controls were identified from the population-representative subset. Two types of polygenic scores were tested for each mental disorder, one trained based on GWAS in EAS and one based on GWAS in both EAS and EUR. SCZ: Schizophrenia. MD: Major depression. GWAS: Genome-wide association studies. EAS: East Asian ancestry. EUR: European ancestry. OR: Odds ratio per standard deviation higher in polygenic scores. CI: Confidence interval.

### Supplementary Table 3. Within-ancestry genetic correlations between mental disorders and other phenotypes.

| **Mental disorder** | **Phenotype** | **Ancestry** | **r_g_** | **SE** | **Z** | ***p*** |
| --- | --- | --- | --- | --- | --- | --- |
| SCZ | MD | EAS | 0.41 | 0.09 | 4.35 | **1.33E-05** |
|  |  | EUR | 0.36 | 0.02 | 21.57 | **3.30E-103** |
|  | BMI | EAS | -0.05 | 0.03 | -1.46 | 1.43E-01 |
|  |  | EUR | -0.11 | 0.01 | -7.25 | **4.20E-13** |
|  | Height | EAS | -0.04 | 0.02 | -1.70 | 9.00E-02 |
|  |  | EUR | -0.01 | 0.01 | -0.77 | 4.40E-01 |
|  | EA | EAS | 0.14 | 0.04 | 3.40 | **7.00E-04** |
|  |  | EUR | 0.03 | 0.02 | 1.96 | **4.96E-02** |
|  | SmkInit | EAS | -0.10 | 0.04 | -2.83 | **4.70E-03** |
|  |  | EUR | 0.17 | 0.02 | 8.88 | **6.86E-19** |
|  | CigDay | EAS | 0.09 | 0.05 | 1.63 | 1.04E-01 |
|  |  | EUR | 0.10 | 0.02 | 4.36 | **1.28E-05** |
| MD | SCZ | EAS | 0.41 | 0.09 | 4.35 | **1.33E-05** |
|  |  | EUR | 0.36 | 0.02 | 21.57 | **3.30E-103** |
|  | BMI | EAS | -0.19 | 0.07 | -2.63 | **8.60E-03** |
|  |  | EUR | 0.17 | 0.02 | 10.84 | **2.16E-27** |
|  | SmkInit | EAS | 0.00 | 0.08 | -0.01 | 9.90E-01 |
|  |  | EUR | 0.38 | 0.02 | 22.43 | **1.95E-111** |
|  | CigDay | EAS | 0.02 | 0.13 | 0.17 | 8.63E-01 |
|  |  | EUR | 0.28 | 0.03 | 9.44 | **3.67E-21** |
|  | Cataract | EAS | -0.10 | 0.18 | -0.58 | 5.62E-01 |
|  |  | EUR | 0.11 | 0.05 | 2.33 | **1.96E-02** |
|  | Diabetes | EAS | -0.08 | 0.06 | -1.35 | 1.77E-01 |
|  |  | EUR | 0.06 | 0.02 | 3.36 | **8.00E-04** |
|  | Stroke | EAS | 0.05 | 0.13 | 0.35 | 7.27E-01 |
|  |  | EUR | 0.89 | 0.03 | 6.54 | **6.24E-11** |
|  | CAD | EAS | -0.04 | 0.10 | -0.39 | 6.96E-01 |
|  |  | EUR | 0.25 | 0.02 | 13.94 | **3.69E-44** |
|  | Lung function | EAS | 0.15 | 0.25 | 0.59 | 5.57E-01 |
|  |  | EUR | 0.00 | 0.01 | 0.44 | 6.60E-01 |
|  | Asthma | EAS | -0.18 | 0.10 | 1.80 | 7.18E-02 |
|  |  | EUR | 0.38 | 0.02 | 17.80 | **7.61E-71** |
|  | RA | EAS | -0.03 | 0.10 | -0.33 | 7.41E-01 |
|  |  | EUR | 0.04 | 0.02 | 1.77 | 7.74E-02 |
|  | PUD | EAS | 0.11 | 0.12 | 0.92 | 3.60E-01 |
|  |  | EUR | 0.16 | 0.03 | 4.98 | **6.51E-07** |
|  | Gallstone | EAS | 0.21 | 0.15 | 1.41 | 1.58E-01 |
|  |  | EUR | 0.29 | 0.03 | 10.52 | **6.64E-26** |

Note. Mental disorder-phenotype pairs that were significant in the phenome-wide association analysis were tested here. Only phenotypes with publicly available GWAS in both EAS and EUR were included. SCZ: Schizophrenia. MD: Major depression. EAS: East Asian ancestry. EUR: European ancestry. BMI: Body mass index. SmkInit: Smoking initiation. CigDay: Cigarettes per day. CAD: Coronary artery disease. RA: Rheumatoid arthritis. PUD: Peptic ulcer disease. SE: Standard error.

### Supplementary Table 4. Cross-ancestry genetic correlations between mental disorders and other phenotypes.

| **Mental disorder (1)** | **Ancestry (1)** | **Phenotype (2)** | **Ancestry (2)** | ***ρ*_ge_** | **SE** | **Z** | ***p*** |
| --- | --- | --- | --- | --- | --- | --- | --- |
| SCZ | EAS | MD | EUR | 0.32 | 0.14 | 3.36 | **7.84E-04** |
|  |  | BMI |  | -0.22 | 0.06 | -3.51 | **4.44E-04** |
|  |  | Height |  | 0.02 | 0.04 | 0.64 | 5.22E-01 |
|  |  | EA |  | 0.15 | 0.06 | 2.55 | **1.09E-02** |
|  |  | SmkInit |  | -0.00 | 0.07 | -0.03 | 9.77E-01 |
|  |  | CigDay |  | -0.01 | 0.06 | -0.18 | 8.60E-01 |
|  | EUR | MD | EAS | 0.32 | 0.14 | 2.25 | **2.43E-02** |
|  |  | BMI |  | -0.12 | 0.05 | -2.61 | **8.95E-03** |
|  |  | Height |  | -0.05 | 0.03 | -1.36 | 1.73E-01 |
|  |  | EA |  | 0.08 | 0.05 | 1.63 | 1.03E-01 |
|  |  | SmkInit |  | 0.05 | 0.05 | 0.94 | 3.50E-01 |
|  |  | CigDay |  | 0.05 | 0.06 | 0.78 | 4.33E-01 |
| MD | EAS | SCZ | EUR | 0.21 | 0.06 | 3.36 | **7.84E-04** |
|  |  | BMI |  | -0.25 | 0.10 | -2.48 | **1.33E-02** |
|  |  | SmkInit |  | -0.01 | 0.10 | -0.09 | 9.32E-01 |
|  |  | CigDay |  | -0.13 | 0.13 | -0.98 | 3.29E-01 |
|  |  | Cataract |  | -0.14 | 0.26 | -0.53 | 5.98E-01 |
|  |  | Diabetes |  | -0.01 | 0.10 | -0.09 | 9.26E-01 |
|  |  | Stroke |  | -0.28 | 0.23 | -1.22 | 2.22E-01 |
|  |  | CAD |  | -0.16 | 0.15 | -1.06 | 2.88E-01 |
|  |  | Lung function |  | -0.08 | 0.06 | -1.25 | 2.12E-01 |
|  |  | Asthma |  | -0.07 | 0.19 | -0.38 | 7.07E-01 |
|  |  | RA |  | NA | NA | NA | NA |
|  |  | PUD |  | 0.04 | 0.20 | 0.21 | 8.34E-01 |
|  |  | Gallstone |  | 0.10 | 0.18 | 0.55 | 5.81E-01 |
|  | EUR | SCZ | EAS | 0.32 | 0.14 | 2.25 | **2.43E-02** |
|  |  | BMI |  | 0.02 | 0.04 | 0.56 | 5.76E-01 |
|  |  | SmkInit |  | 0.30 | 0.04 | 6.86 | **6.82E-12** |
|  |  | CigDay |  | 0.24 | 0.05 | 5.19 | **2.05E-07** |
|  |  | Cataract |  | 0.10 | 0.07 | 1.40 | 1.60E-01 |
|  |  | Diabetes |  | 0.07 | 0.03 | 2.35 | **1.88E-02** |
|  |  | Stroke |  | 0.07 | 0.06 | 1.08 | 2.81E-01 |
|  |  | CAD |  | 0.06 | 0.04 | 1.47 | 1.43E-01 |
|  |  | Lung function |  | -0.09 | 0.11 | -0.81 | 4.18E-01 |
|  |  | Asthma |  | 0.10 | 0.05 | -1.97 | **4.89E-02** |
|  |  | RA |  | -0.05 | 0.05 | -1.07 | 2.84E-01 |
|  |  | PUD |  | 0.16 | 0.06 | 2.79 | **5.27E-03** |
|  |  | Gallstone |  | 0.12 | 0.07 | 1.68 | 9.25E-02 |

Note. Mental disorder-phenotype pairs that were significant in the phenome-wide association analysis were tested here. Only phenotypes with publicly available GWAS in both EAS and EUR were included. Mental disorders (1) in one ancestry were tested against phenotypes (2) in the other ancestry. NA represents unreliable estimates that had SE > 0.3. SCZ: Schizophrenia. MD: Major depression. EAS: East Asian ancestry. EUR: European ancestry. BMI: Body mass index. SmkInit: Smoking initiation. CigDay: Cigarettes per day. CAD: Coronary artery disease. RA: Rheumatoid arthritis. PUD: Peptic ulcer disease. SE: Standard error.

### Supplementary Table 5. Phenotypic correlations between mental disorders and phenotypes in the UK Biobank.

|  | BMI | SmkInit | CigDay | CanUD | EA | Income | SCZ | MD |
| --- | --- | --- | --- | --- | --- | --- | --- | --- |
| BMI | 1.000 | 0.034 | 0.092 | -0.020 | -0.105 | -0.095 | 0.012 | 0.063 |
| SmkInit | 0.034 | 1.000 | 0.000 | 0.122 | -0.064 | -0.046 | 0.012 | 0.038 |
| CigDay | 0.092 | 0.000 | 1.000 | -0.098 | -0.077 | -0.106 | 0.073 | 0.070 |
| CanUD | -0.020 | 0.122 | -0.098 | 1.000 | 0.027 | 0.012 | 0.010 | 0.025 |
| EA | -0.105 | -0.064 | -0.077 | 0.027 | 1.000 | 0.331 | -0.001 | -0.038 |
| Income | -0.095 | -0.046 | -0.106 | 0.012 | 0.331 | 1.000 | -0.049 | -0.120 |
| SCZ | 0.012 | 0.012 | 0.073 | 0.010 | -0.001 | -0.049 | 1.000 | 0.064 |
| MD | 0.063 | 0.038 | 0.070 | 0.025 | -0.038 | -0.120 | 0.064 | 1.000 |

Note. Numbers represent Spearman's rank correlation coefficient (*ρ*). Heavy cannabis use (lifetime cannabis use > 100 times) was used as a proxy for CanUD in the UK Biobank. BMI: Body mass index. BMI: Body mass index. SmkInit: Smoking initiation. CigDay: Cigarettes per day. CanUD: Cannabis use disorder. EA: Educational attainment. SCZ: Schizophrenia. MD: Major depression.

### Supplementary Figure 1. Performance of schizophrenia polygenic scores in CKB.

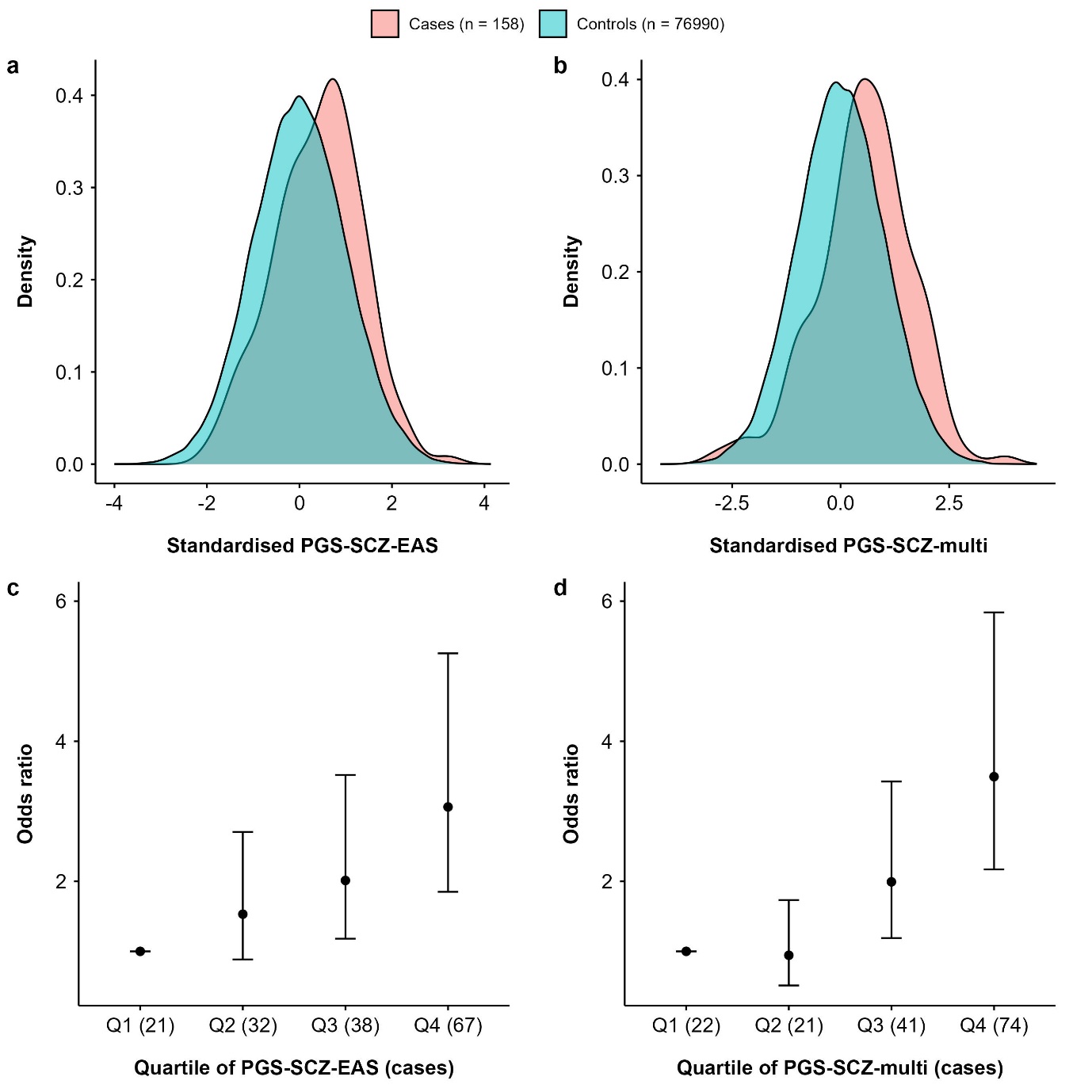

Note. a, Distribution of PGS-SCZ-EAS. b, Distribution of PGS-SCZ-multi. c, Associations between PGS-SCZ-EAS and SCZ by quartile. d, Associations between PGS-SCZ-multi and SCZ by PGS quartile. Cases of SCZ were identified from the overall dataset, while controls were identified from the population-representative subset. Two types of PGS tested, one based on GWAS in EAS and one based on GWAS in both EAS and EUR. PGS: Polygenic score. SCZ: Schizophrenia. EAS: East Asian ancestry. EUR: European ancestry.

### Supplementary Figure 2. Performance of major depression polygenic scores in CKB.

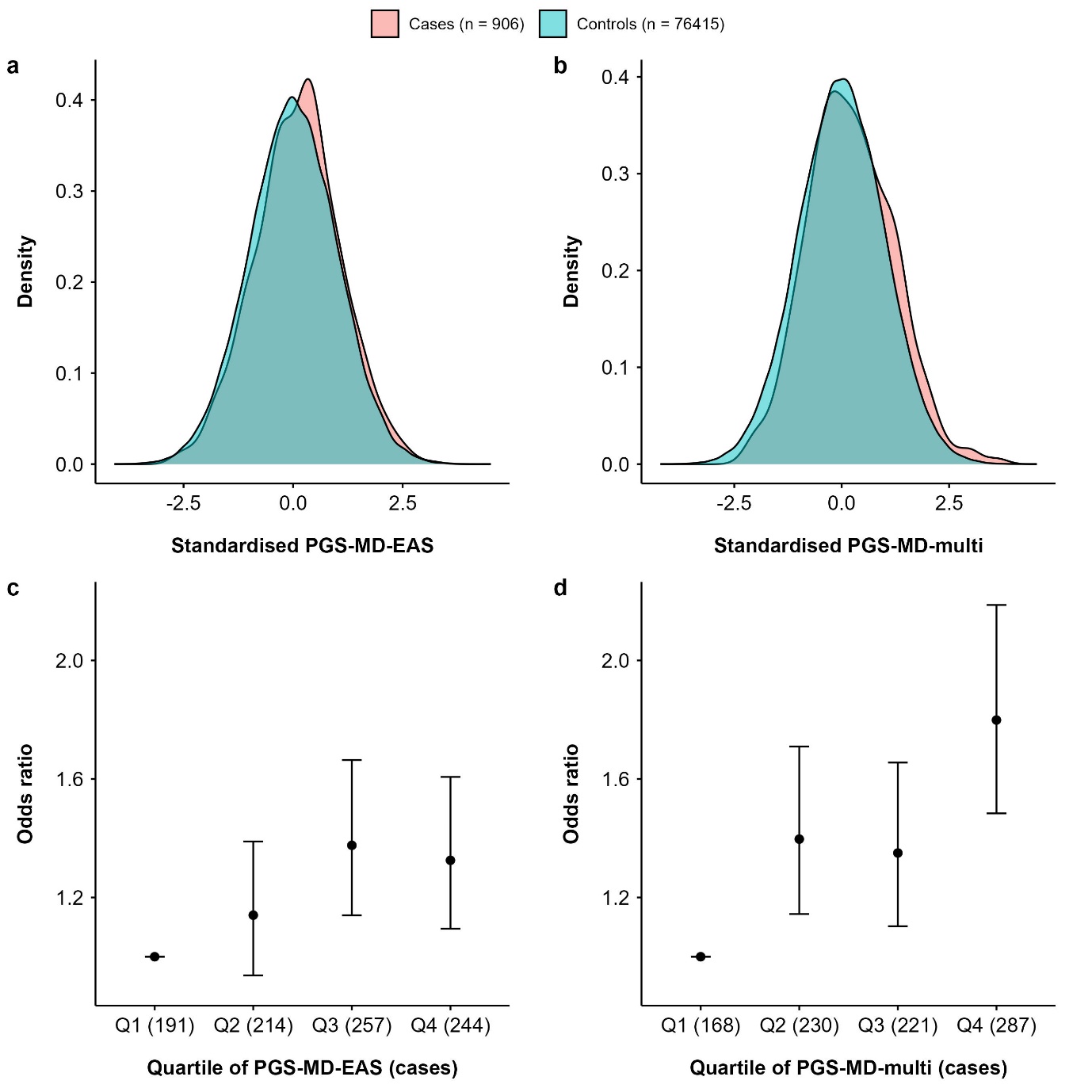

Note. a, Distribution of PGS-MD-EAS. b, Distribution of PGS-MD-multi. c, Associations between PGS-MD-EAS and MD by quartile. d, Associations between PGS-MD-multi and MD by PGS quartile. Cases of MD were identified from the overall dataset, while controls were identified from the population-representative subset. Two types of PGS tested, one based on GWAS in EAS and one based on GWAS in both EAS and EUR. PGS: Polygenic score. MD: Major depression. EAS: East Asian ancestry. EUR: European ancestry.

### Supplementary Figure 3. Performance of schizophrenia polygenic scores among females in CKB.

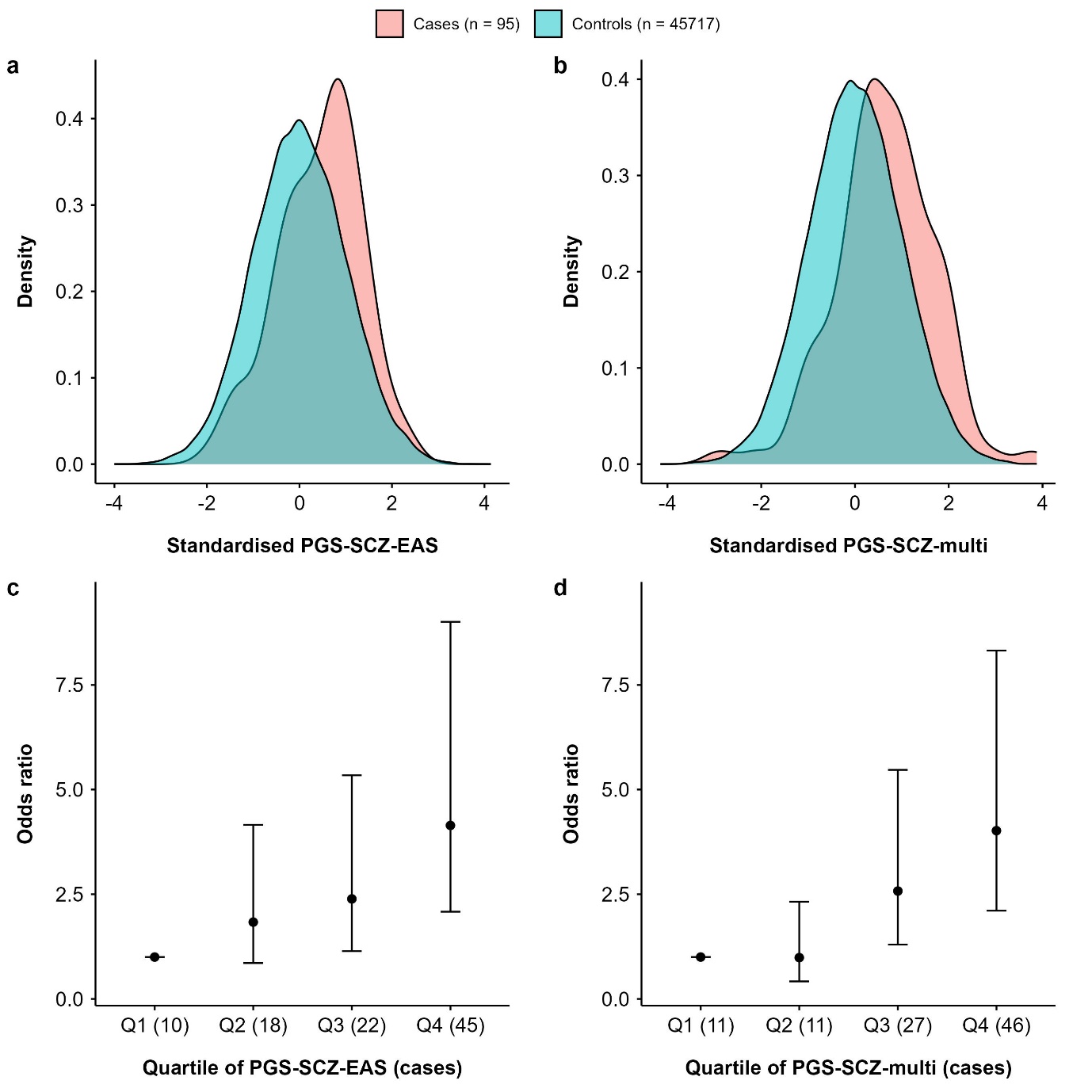

Note. a, Distribution of PGS-SCZ-EAS. b, Distribution of PGS-SCZ-multi. c, Associations between PGS-SCZ-EAS and SCZ by quartile. d, Associations between PGS-SCZ-multi and SCZ by PGS quartile. Cases of SCZ were identified from the overall dataset, while controls were identified from the population-representative subset. Two types of PGS tested, one based on GWAS in EAS and one based on GWAS in both EAS and EUR. PGS: Polygenic score. SCZ: Schizophrenia. EAS: East Asian ancestry. EUR: European ancestry.

### Supplementary Figure 4. Performance of major depression polygenic scores among females in CKB.

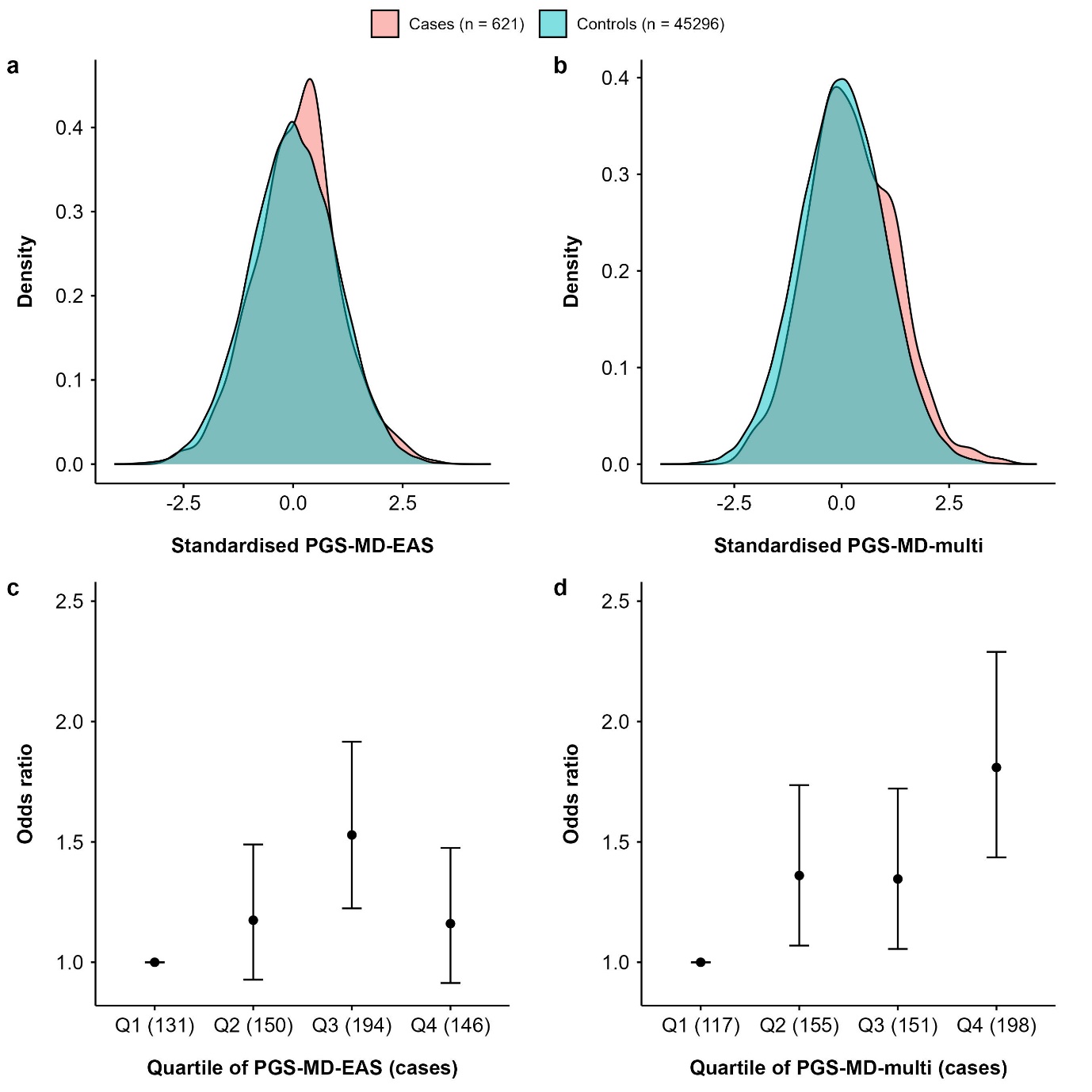

Note. a, Distribution of PGS-MD-EAS. b, Distribution of PGS-MD-multi. c, Associations between PGS-MD-EAS and MD by quartile. d, Associations between PGS-MD-multi and MD by PGS quartile. Cases of MD were identified from the overall dataset, while controls were identified from the population-representative subset. Two types of PGS tested, one based on GWAS in EAS and one based on GWAS in both EAS and EUR. PGS: Polygenic score. MD: Major depression. EAS: East Asian ancestry. EUR: European ancestry.

### Supplementary Figure 5. Performance of schizophrenia polygenic scores among males in CKB.

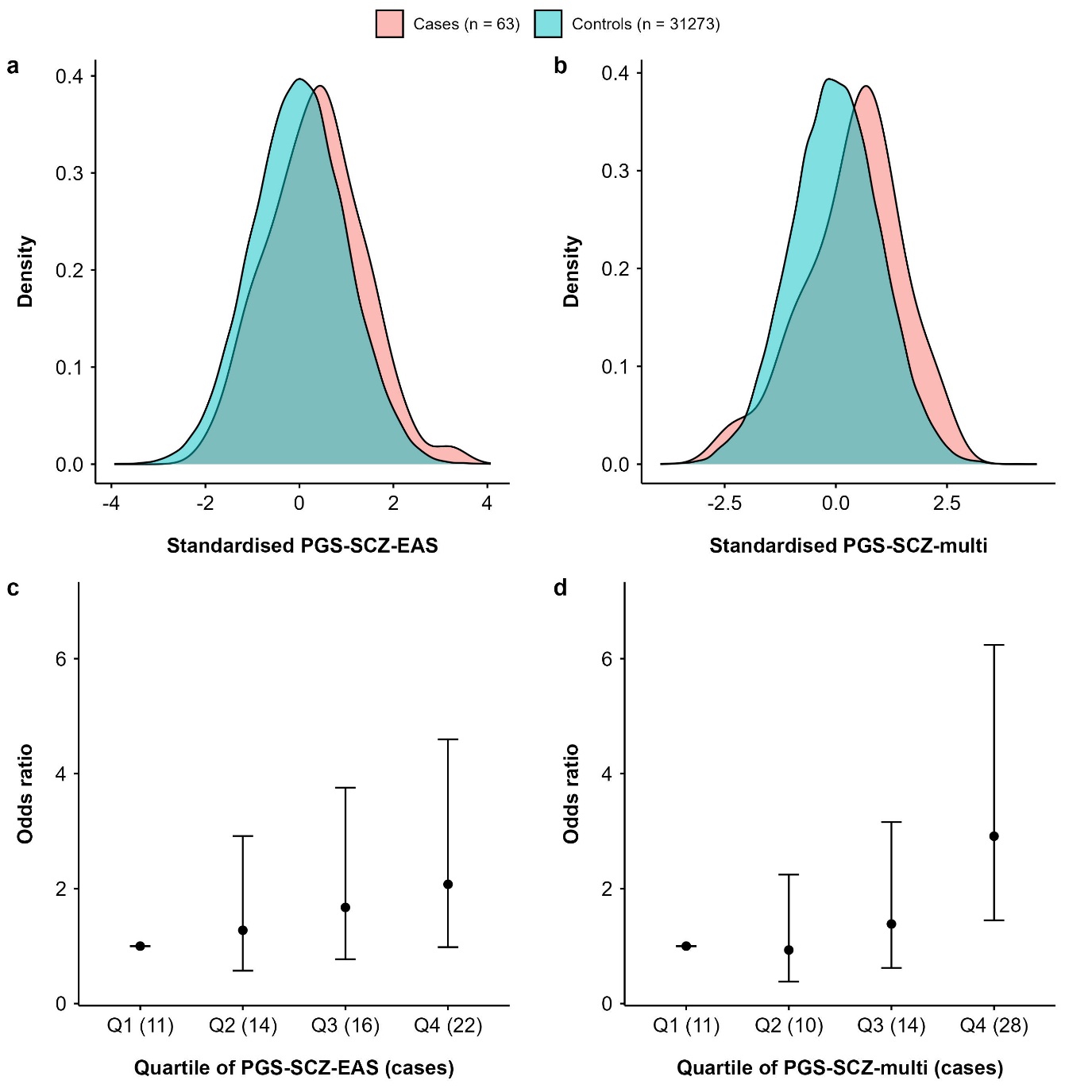

Note. a, Distribution of PGS-SCZ-EAS. b, Distribution of PGS-SCZ-multi. c, Associations between PGS-SCZ-EAS and SCZ by quartile. d, Associations between PGS-SCZ-multi and SCZ by PGS quartile. Cases of SCZ were identified from the overall dataset, while controls were identified from the population-representative subset. Two types of PGS tested, one based on GWAS in EAS and one based on GWAS in both EAS and EUR. PGS: Polygenic score. SCZ: Schizophrenia. EAS: East Asian ancestry. EUR: European ancestry.

### Supplementary Figure 6. Performance of major depression polygenic scores among males in CKB.

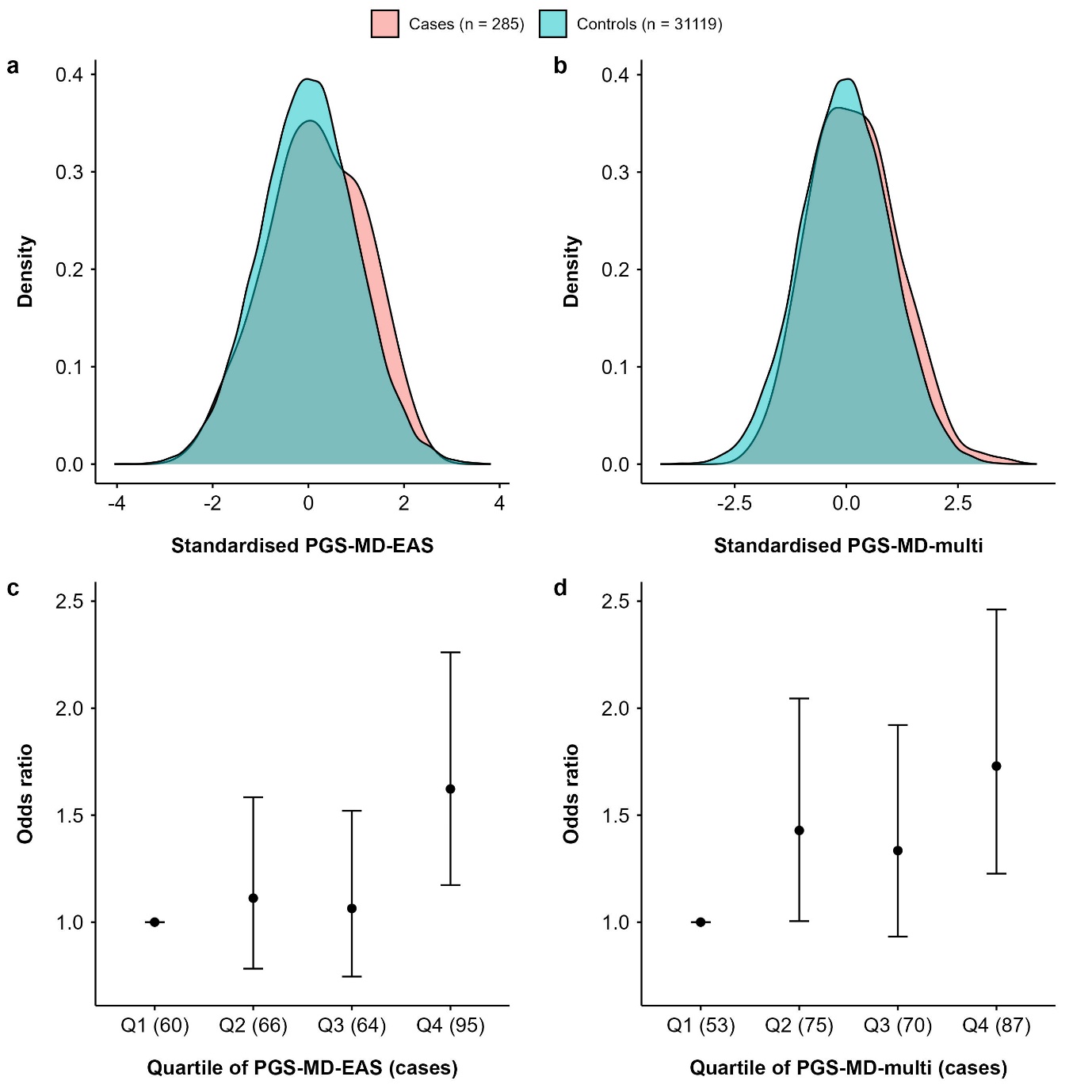

Note. a, Distribution of PGS-MD-EAS. b, Distribution of PGS-MD-multi. c, Associations between PGS-MD-EAS and MD by quartile. d, Associations between PGS-MD-multi and MD by PGS quartile. Cases of MD were identified from the overall dataset, while controls were identified from the population-representative subset. Two types of PGS tested, one based on GWAS in EAS and one based on GWAS in both EAS and EUR. PGS: Polygenic score. MD: Major depression. EAS: East Asian ancestry. EUR: European ancestry.

### Supplementary Figure 7. Phenome-wide associations with polygenic scores for schizophrenia and major depression among females in CKB.

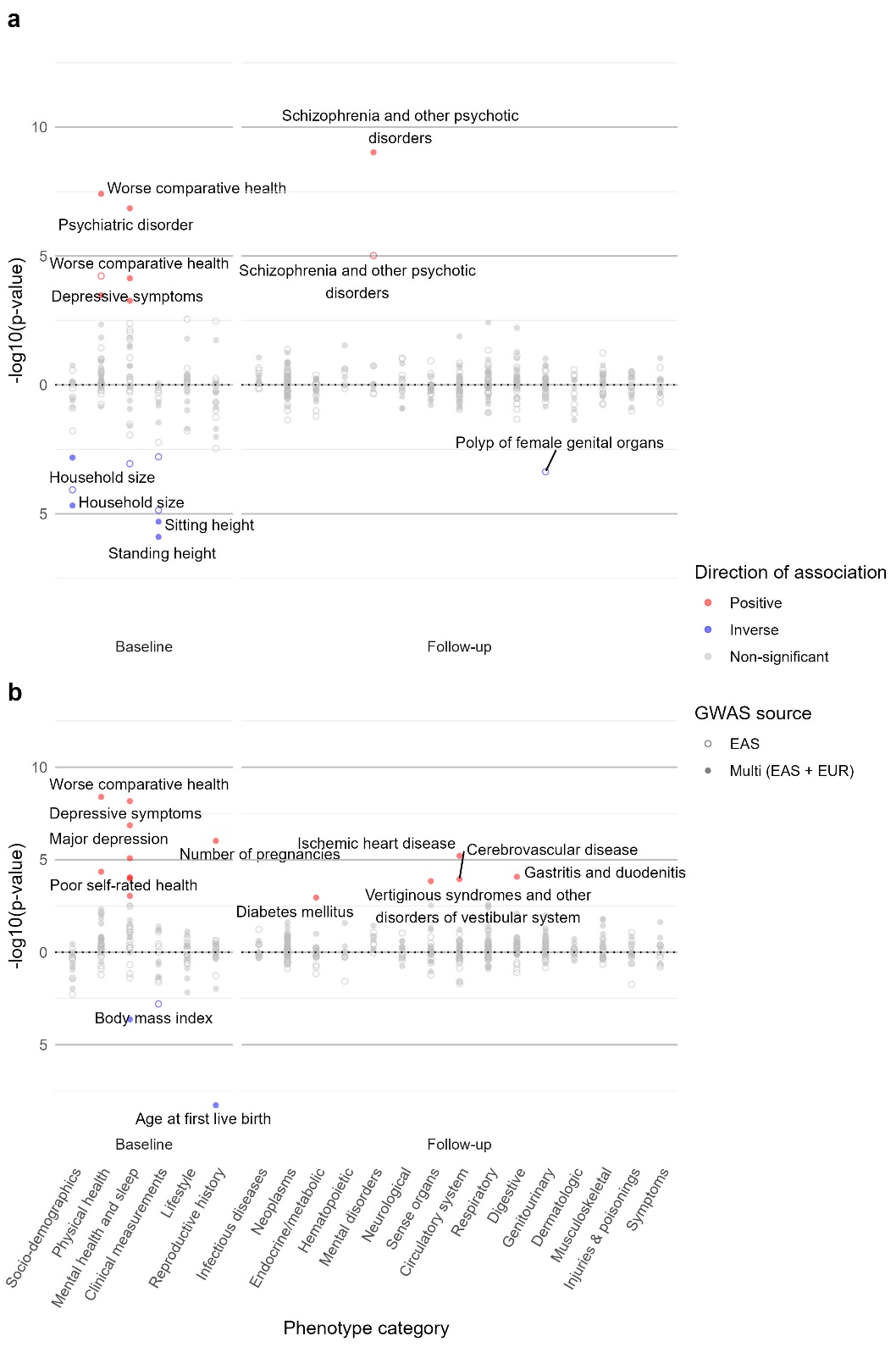

Note. a, Results of polygenic scores for schizophrenia. b, Results of polygenic scores for major depression. A total of 250 phenotypes (67 at baseline and 183 at follow-up) were tested. The shape of dots indicates the GWAS source, while the colour of dots indicates the direction of association. Results were corrected for multiple testing with a false discovery rate = 0.05. The top two most significant associations in each phenotype category are labelled. PGS: Polygenic score. SCZ: Schizophrenia. MD: Major depression. GWAS: Genome-wide association studies. EAS: East Asian ancestry. EUR: European ancestry.

### Supplementary Figure 8. Phenome-wide associations with polygenic scores for schizophrenia and major depression among males in CKB.

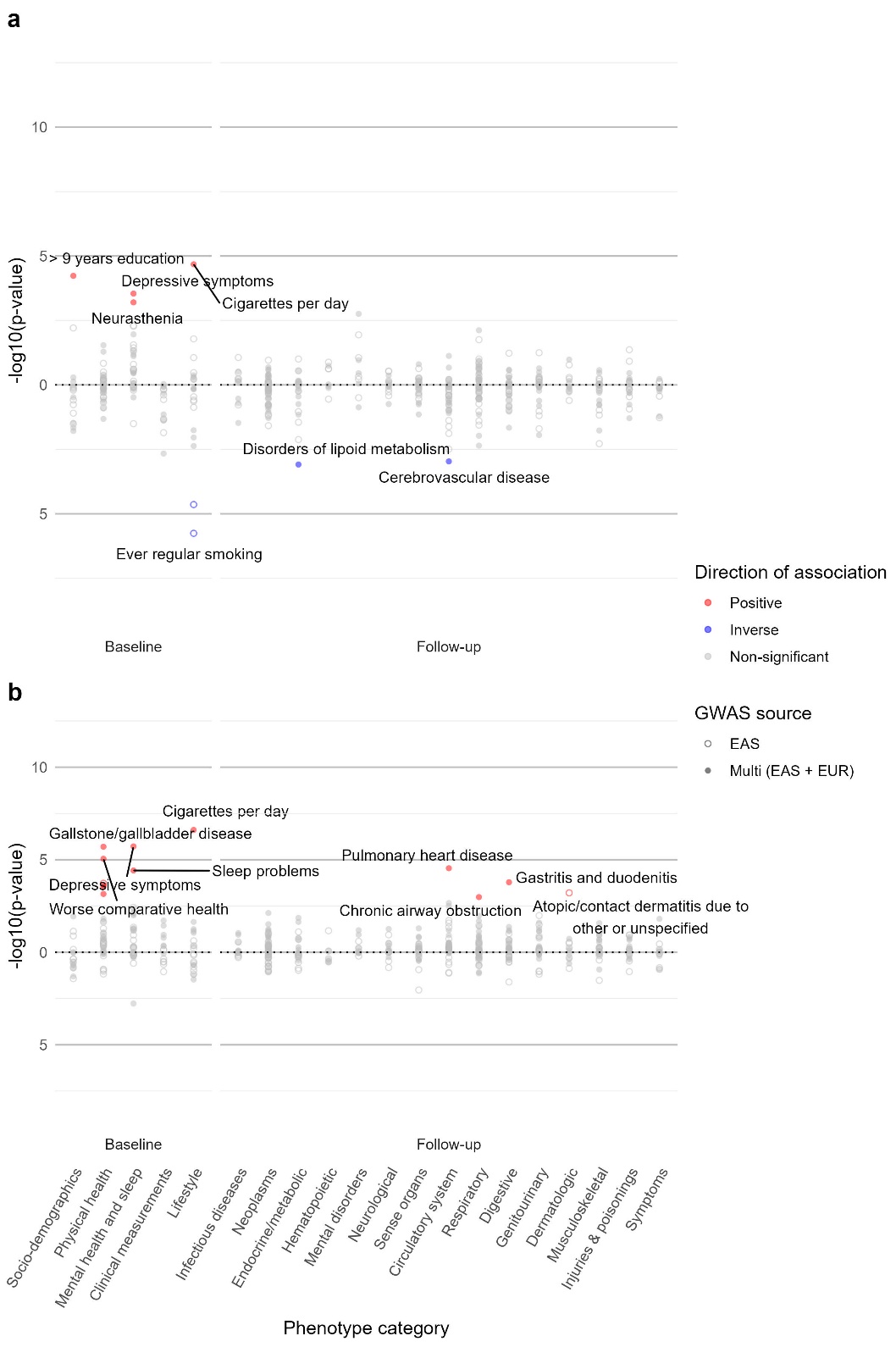

Note. a, Results of polygenic scores for schizophrenia. b, Results of polygenic scores for major depression. A total of 232 phenotypes (57 at baseline and 175 at follow-up) were tested. The shape of dots indicates the GWAS source, while the colour of dots indicates the direction of association. Results were corrected for multiple testing with a false discovery rate = 0.05. The top two most significant associations in each phenotype category are labelled. PGS: Polygenic score. SCZ: Schizophrenia. MD: Major depression. GWAS: Genome-wide association studies. EAS: East Asian ancestry. EUR: European ancestry.

### Supplementary Figure 9. Cross-ancestry genetic correlations between schizophrenia/major depression and related phenotypes.

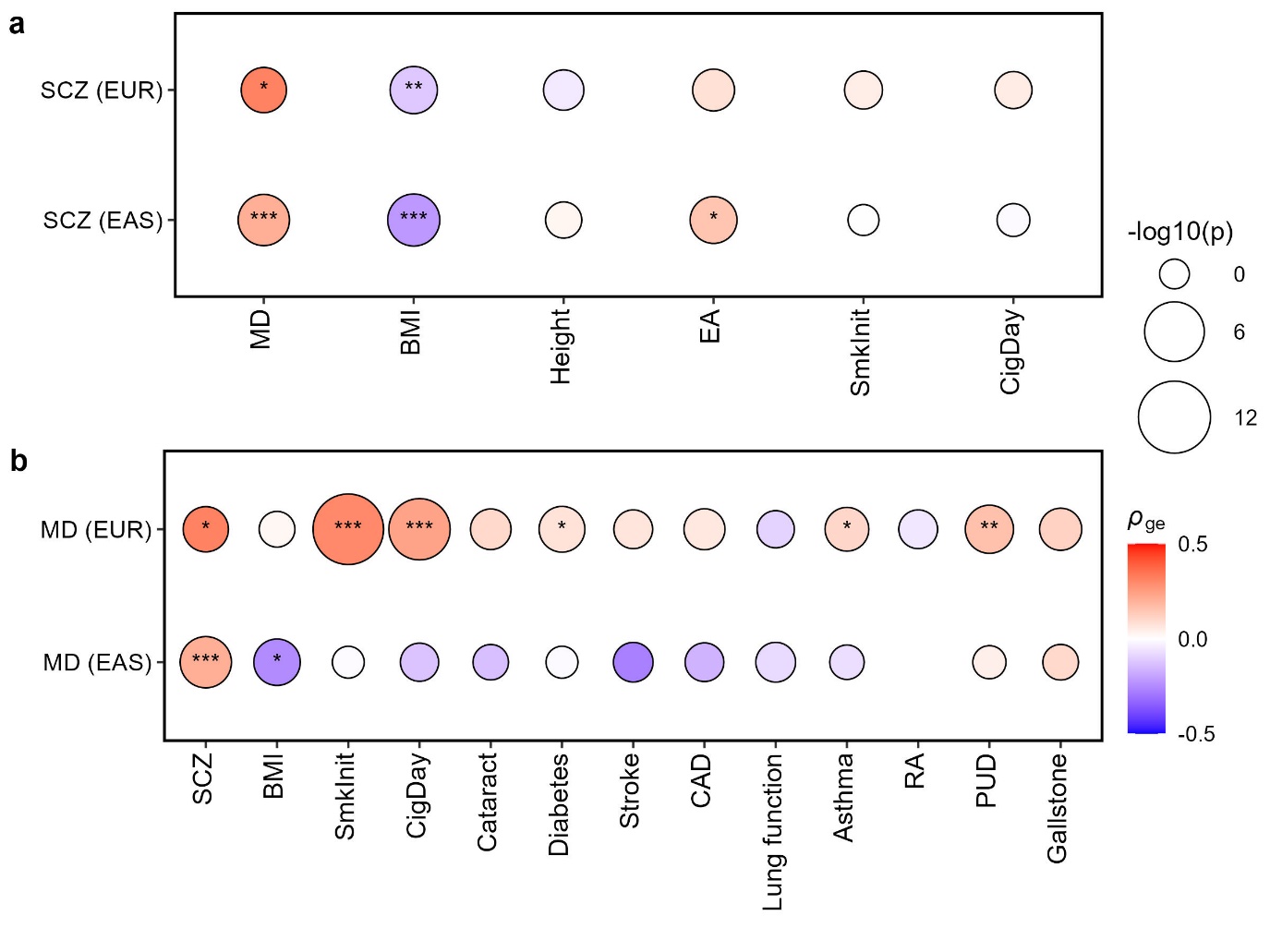

Note. a, Genetic correlations with SCZ. b, Genetic correlations with MD. Cross-ancestry genetic effect correlations (*ρ*_ge_) were tested by Popcorn, using GWAS on SCZ/MD in one ancestry and GWAS on phenotypes in the other ancestry. Mental disorder-phenotype pairs that were significant in the phenome-wide association analysis were tested here. Only phenotypes with publicly available GWAS in both EAS and EUR were included. The colour of the circles indicates the correlation coefficient. The size of the circles is scaled to -log10(p-value). *: *p* < 0.05; **: *p* < 0.01; ***: *p* < 0.001. Only estimates with SE < 0.3 are shown here. SCZ: Schizophrenia. MD: Major depression. EAS: East Asian ancestry. EUR: European ancestry. BMI: Body mass index. SmkInit: Smoking initiation. CigDay: Cigarettes per day. CAD: Coronary artery disease. RA: Rheumatoid arthritis. PUD: Peptic ulcer disease. SE: Standard error.

### Supplementary Figure 10. Bi-directional Mendelian Randomisation between schizophrenia/major depression and other phenotypes with a more stringent threshold.

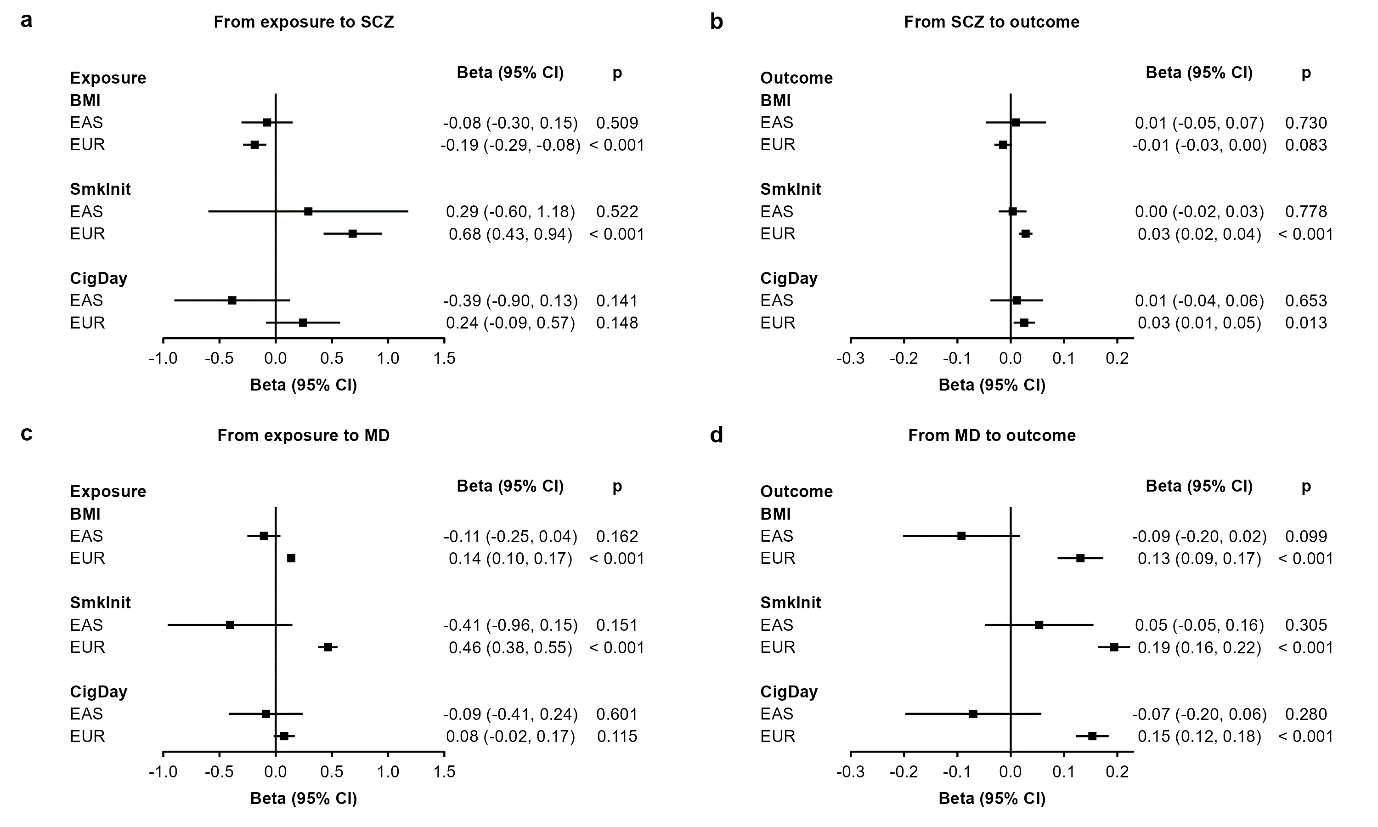

Note. a, From exposure to SCZ. b, From SCZ to outcome. c, From exposure to MD. d, From MD to outcome. Results shown here are based on estimates from the Wald ratio test (MD as exposure) and the inverse variance weighted method (all other exposures). Genetic instruments were selected based on *p* < 5 × 10^-8^ in both EAS and EUR after clumping. SCZ: Schizophrenia. MD: Major depression. EAS: East Asian ancestry. EUR: European ancestry. BMI: Body mass index. SmkInit: Smoking initiation. CigDay: Cigarettes per day. CI: Confidence interval.

### Supplementary Figure 11. Selection of cases and controls in PheWAS.

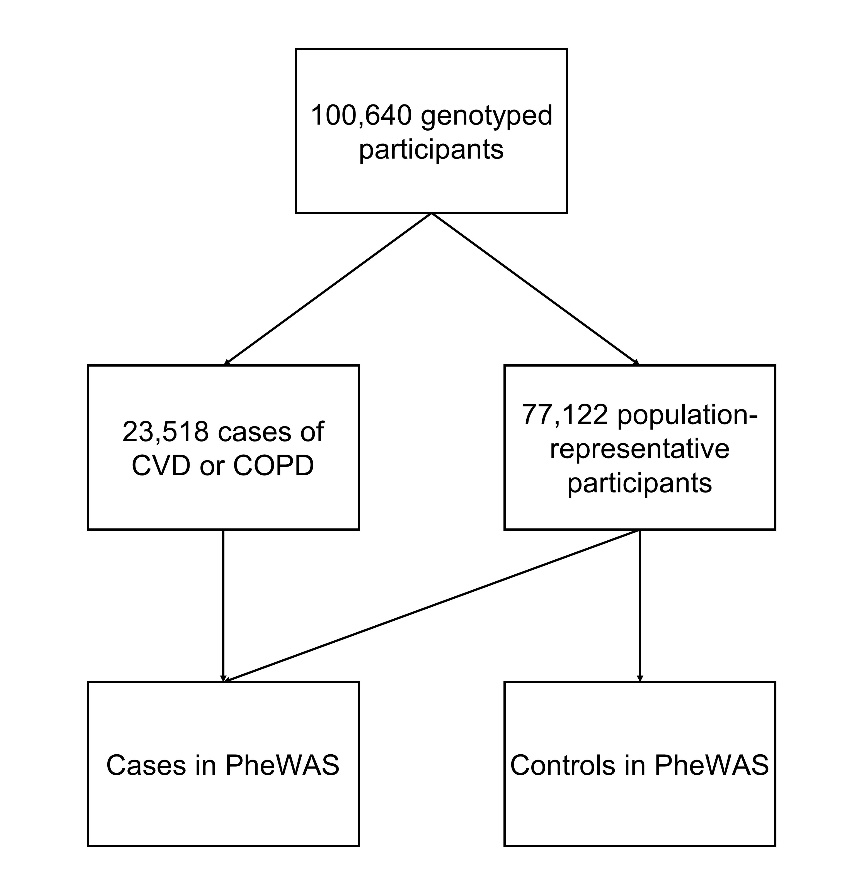

Note. In PheWAS, cases of all disease-related phenotypes were identified from the overall dataset (including both selected CVD/COPD cases and population-representative participants), while controls were only identified from the population-representative subset. CVD: Cardiovascular disease. COPD: Chronic obstructive pulmonary disease.
